# Internal and External Validation of Comprehensive High-Frequency Activity Biomarkers for Epilepsy Surgery

**DOI:** 10.64898/2025.12.17.25342490

**Authors:** Keisuke Hatano, Naoto Kuroda, Hiroshi Uda, Kazuki Sakakura, Michael J. Cools, Aimee F. Luat, Shin-Ichiro Osawa, Hitoshi Nemoto, Kazushi Ukishiro, Hidenori Endo, Nobukazu Nakasato, Yutaro Takayama, Keiya Iijima, Masaki Iwasaki, Eishi Asano

**Author notes:** **Correspondence to:** Eishi Asano, M.D., Ph.D., M.S. (C.R.D.S.A.) Full address: Division of Pediatric Neurology, Children’s Hospital of Michigan, Wayne State University. 3901 Beaubien St., Detroit, MI, 48201, USA.

## Abstract

Although epilepsy surgery studies have proposed intracranial EEG–derived biomarkers for localizing seizure onset and anticipating postoperative outcomes, evaluation has often been limited to derivation cohorts using internal cross-validation. An influential notion holds that neurons distributed within the seizure onset zone (SOZ) frequently generate high-frequency activity (HFA) and that resection of such sites is associated with favorable postoperative seizure control. However, the extent to which these prediction models generalize to independent patient populations—and across diverse underlying etiologies—has remained largely untested. In this international, multi-center study of drug-resistant focal epilepsy, we retrospectively quantified HFA occurrence rates together with a comprehensive set of morphological features and integrated these metrics into predictive models for SOZ localization and postoperative seizure outcome. We then assessed model performance in fully independent datasets—a temporal external cohort and two geographical external cohorts—each entirely separate from the derivation cohort. In total, 5,142,891 HFA events observed across 22,939 electrodes from 233 patients were analyzed. Among the model inputs, HFA rate, spectral entropy, and power emerged as the most influential features for accurate SOZ classification. The model reliably identified clinician-defined SOZ sites across centers, achieving areas under the curve (AUCs) of up to 0.85 in the derivation cohort using 10-fold cross-validation, up to 0.86 in the temporal external cohort, and up to 0.75 in the geographical external cohorts. Within the derivation cohort, the model predicted postoperative seizure freedom with an AUC of up to 0.70. In contrast, postoperative seizure outcomes could not be predicted reliably across the external cohorts. Specifically, among external-cohort patients with MRI non-lesional epilepsy, postoperative seizure freedom was predicted with an AUC of up to 0.73, whereas performance declined to 0.46 or below among patients with encephalomalacia, an etiology characterized by chronic parenchymal damage and marked neuronal loss. Together, integration of HFA occurrence rates with morphological features yields an SOZ-localization biomarker with cross-center generalizability, whereas postoperative outcome prediction remains highly dependent on underlying etiology. Notably, a surgical strategy that prioritizes resection of HFA-involved areas does not appear to be applicable to patients with encephalomalacia and may be ineffective or even counterproductive in this population. A total of 97.86 GB of iEEG data are publicly available to facilitate external validation by the epilepsy surgery research community and the development of improved biomarkers for epileptogenic zone localization.

## Introduction

The ultimate goal of resective surgery for drug-resistant focal epilepsy is to achieve long-term seizure freedom^1,2^. Conventional surgical planning frequently relies on intracranial EEG (iEEG) monitoring to capture habitual seizures, delineate the seizure onset zone (SOZ), and guide cortical resection in conjunction with MRI findings^3^. Although the SOZ is not a perfect construct, it often serves as a practical target for surgical removal^4–7^. Nevertheless, this approach faces a critical limitation: habitual seizures may not occur promptly after electrode implantation, imposing physical and psychological burdens on patients. Accordingly, researchers have sought iEEG biomarkers that can be measured over shorter recordings and localize regions corresponding to the SOZ. A biomarker that also improves prediction of postoperative seizure outcomes would provide even greater clinical value. Importantly, a biomarker validated only within an internal dataset is limited to explaining variance in past outcomes from the same institution. In contrast, when its performance is confirmed in an independently acquired external dataset, the biomarker can be considered generalizable and thus more likely to translate to real-world clinical practice.

In this retrospective, multicenter observational study, we investigated the utility of interictal high-frequency activity (HFA) as a candidate iEEG biomarker for localizing the SOZ and predicting postoperative seizure outcomes. HFA was defined as a brief event exceeding 80 Hz that stands out distinctly from the surrounding background activity. To ensure objectivity and reproducibility, HFA was detected using four widely adopted automatic algorithms^8–11^, focusing on epochs of non–REM sleep. Prior studies have shown that the SOZ tends to generate HFA more frequently than the remaining areas^12^. A meta-analysis and a registered clinical trial suggest that resections encompassing regions rich in interictal HFA, if observed, are associated with improved postoperative outcomes^13,14^.

Mechanistically, HFA represents a heterogeneous phenomenon: some events manifest as discrete oscillatory bursts, referred to as high-frequency oscillations (HFOs); others correspond to interictal spikes with broadband high-frequency augmentation; and some occur as spike–HFO complexes^15–17^. The clinical relevance of these subtypes remains a matter of debate. For example, a randomized trial comparing resections guided by intraoperative HFA versus interictal spikes demonstrated comparable or slightly better outcomes when guided by spikes^18^. Moreover, independent research groups have shown that resection of cortical sites generating spike–HFO coupling is associated with postoperative seizure freedom^17,19^. Collectively, these findings have led to our hypothesis that the diagnostic utility of HFA depends not only on its occurrence rate but also on its morphological characteristics.

We therefore hypothesized that moving beyond occurrence rates—by integrating *quantifiable* morphological features such as power, frequency, duration, and spectral entropy along with anatomical information and patient age^20–23^—would provide a more robust biomarker of the SOZ and improve the prediction of surgical outcomes. Spectral entropy provides a quantitative measure of an event’s spectral structure: lower values indicate narrow-band oscillatory HFA, likely representing HFOs, whereas higher values reflect broadband HFA characteristic of spikes^24^. Initially, we tested whether an eXtreme Gradient Boosting (XGBoost) model^25^ that combined HFA rate, morphological and anatomical features, and patient age could retrospectively account for both SOZ localization and postoperative seizure freedom in a large derivation cohort using cross-validation (internal validation). We then examined whether the SOZ classification model derived from historical data could accurately predict SOZ sites and postoperative outcomes in a newly acquired, independent cohort from the same center (*temporal* external validation). Finally, we evaluated whether this model generalized to datasets collected by independent investigators at two additional centers (*geographical* external validation).

Drug-resistant focal epilepsy arises from heterogeneous etiologies. Accordingly, within external validation cohorts, we sought to determine which MRI-defined etiologies were associated with variability in model performance. Because pathological HFA is thought to arise from *neural* populations distributed within the epileptogenic zone^26–28^, we evaluated model performance separately across MRI-defined etiologies. Specifically, we examined patients with MRI-nonlesional epilepsy and those with encephalomalacia (often referred to as ulegyria), a form of chronic parenchymal injury accompanied by marked neuronal loss that is considered to constitute the epileptogenic zone^29,30^. We additionally assessed model performance in patients with tumors and focal cortical dysplasia, given that, in tumor-related epilepsy, the epileptogenic zone has been suggested to involve the peritumoral cortex, whereas focal cortical dysplasia is widely recognized as intrinsically epileptogenic^31,32^.

## Materials and methods

### Patient cohorts

The inclusion criteria were as follows: (1) patients with drug-resistant focal epilepsy who underwent curative resective surgery following extraoperative iEEG monitoring; (2) availability of postoperative follow-up data for at least one year; and (3) interictal iEEG recordings obtained during non-REM sleep, recorded at least two hours apart from seizure events. The exclusion criteria were as follows: (1) hemispherotomy or hemispherectomy; (2) suspected malignant brain tumor based on preoperative MRI findings or medical history; (3) prior resective epilepsy surgery; and (4) severe brain malformations precluding accurate identification of the central or lateral sulcus.

The cohort of patients who underwent iEEG recording at the Detroit Medical Center (Detroit, USA) between January 2007 and August 2016 was designated as the derivation cohort and used for model training and internal cross-validation. Patients who underwent iEEG recording at the same center between September 2016 and May 2022 constituted the *temporal* external validation cohort. In addition, patients who underwent iEEG recording at Tohoku University Hospital (Sendai, Japan) between January 2011 and December 2015, as well as those studied at the National Center of Neurology and Psychiatry (NCNP; Tokyo, Japan) between February 2017 and July 2019, constituted the *geographical* external validation cohorts. The study protocol was approved by the Institutional Review Boards of Wayne State University (#048404MP2E), Tohoku University (#2023-1-1114-1), and the National Center of Neurology and Psychiatry (#A2021-050). Written informed consent was obtained from all participants or from the legal guardians of pediatric patients.

### iEEG

The general procedures have been described previously^33,34^. Platinum subdural electrodes (3 mm diameter; 10 mm interelectrode distance) and/or depth electrodes (2 mm in length; 3.5–4.5 mm interelectrode distance) were implanted via craniotomy or stereotactically to delineate the boundaries of the presumed epileptogenic zone and functionally important cortical areas. In the epilepsy monitoring unit, iEEG signals were recorded simultaneously with video for 2–14 days using the Nihon Kohden Digital System (Nihon Kohden, Tokyo, Japan) with a sampling rate of 1,000 Hz and an amplifier band-pass filter of 0.016–300 Hz. In general, antiseizure medications (ASMs) were tapered or discontinued during monitoring and resumed after localization of the SOZ. The SOZ was defined as the electrode sites showing the earliest unequivocal, sustained rhythmic or repetitive discharges that were not attributable to state changes and that preceded the habitual seizure symptoms. The SOZ was prospectively determined by board-certified clinical neurophysiologists or epileptologists at each institution.

### HFA characterization

We extracted the earliest, least-artifactual iEEG data (20-minute epoch for the Detroit Medical Center cohort; 5-minute epoch for the geographically external validation cohorts) recorded during non-REM sleep, encompassing both stage 2 and slow-wave sleep^23^. The selected data were required to be at least 2 hours away from seizure events. No notch filter was applied. Channels exhibiting artifacts or located within white matter or outside the brain were excluded from further analysis.

Four established algorithms implemented in RIPPLELAB^35^ (https://github.com/BSP-Uniandes/RIPPLELAB) were employed for automatic HFA detection: Short Time Energy^8^ (STE), Short Line Length^9^ (SLL), Hilbert^10^ (HIL), and Montreal Neurological Institute^11^ (MNI). Each algorithm first band-pass filters the iEEG signal (80–500 Hz) and then identifies transient high-frequency events using detector-specific amplitude or energy criteria as originally defined in the cited studies. Default RIPPLELAB parameters were used (see Kuroda et al., 2021^36^ for details).

We quantified the duration (ms), maximum spectral power (µV²), frequency band at maximum spectral power (Hz), and normalized spectral entropy as morphological features of each HFA event. A common average montage was applied to subdural electrode channels, whereas a bipolar montage was used for depth electrode channels to minimize far-field effects and reduce volume conduction^37,38^. Each depth contact, assessed with a bipolar montage, was assigned the mean of two values derived from its adjacent electrode pairs. RIPPLELAB automatically computed HFA event duration, whereas the remaining features were derived through time–frequency analysis using the multitaper method implemented in the FieldTrip toolbox^39,40^ (https://www.fieldtriptoolbox.org/). The time–frequency representation was generated using 200-ms time windows and 5-Hz frequency bins, sliding by 1 ms. The maximum spectral power and its corresponding frequency (i.e., the frequency band in which the power peaked) were extracted as morphological features. Spectral entropy was calculated from the normalized spectral power distribution during each HFA to quantify spectral concentration. Spectral entropy (*H*), equivalent to Shannon entropy, was defined as^24^:

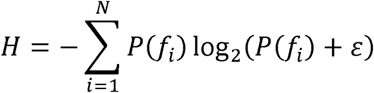

where *N* is the number of 5-Hz frequency bins, *P*(*f_i_*) is the normalized power at frequency *f_i_*, and ε is a small constant added to avoid logarithm of zero. To enable comparison across different spectral resolutions, the entropy was normalized by the maximum possible entropy,

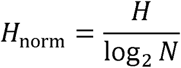

Accordingly, normalized spectral entropy (*H*_norm_ ) ranges from 0 to 1, with lower values indicating HFAs characterized by power augmentation concentrated within a narrow frequency band, and higher values indicating broadband power increases resembling interictal spike discharges (**Figure S1**).

## Imaging

Three-dimensional cortical surface images were reconstructed from preoperative 3T T1-weighted spoiled gradient–echo volumetric MRI using the FreeSurfer software package (http://surfer.nmr.mgh.harvard.edu) for patients aged two years or older, and the Infant FreeSurfer package (https://surfer.nmr.mgh.harvard.edu/fswiki/infantFS) for patients younger than two years^23^. When the pial surface could not be delineated because of insufficient cerebral myelination, a board-certified neurosurgeon (K.S. or K.H.) accurately delineated it using the *Control Point* function (https://surfer.nmr.mgh.harvard.edu/fswiki/FsTutorial/ControlPoints_freeview/).

Electrode sites were displayed on the pial surface by co-registering preoperative MRI with post-implant CT images using the FieldTrip toolbox^41^. At least two board-certified neurosurgeons (K.H., N.K., and K.S.) visually inspected intraoperative photographs and CT images to confirm the spatial accuracy of electrode locations co-registered to the MRI-derived cortical surface. Depth electrodes located more than 2 mm from the nearest cortex were considered unattached and excluded from further analysis. Each cortical electrode was assigned a coordinate on the FreeSurfer surface^23,42^. Each cortical electrode was mapped onto the FreeSurfer cortical surface and assigned to one of 12 anatomical regions (frontal, central, parietal, occipital, temporal, or insular cortices in both hemispheres; **Figure 1A**).

**Figure 1.**
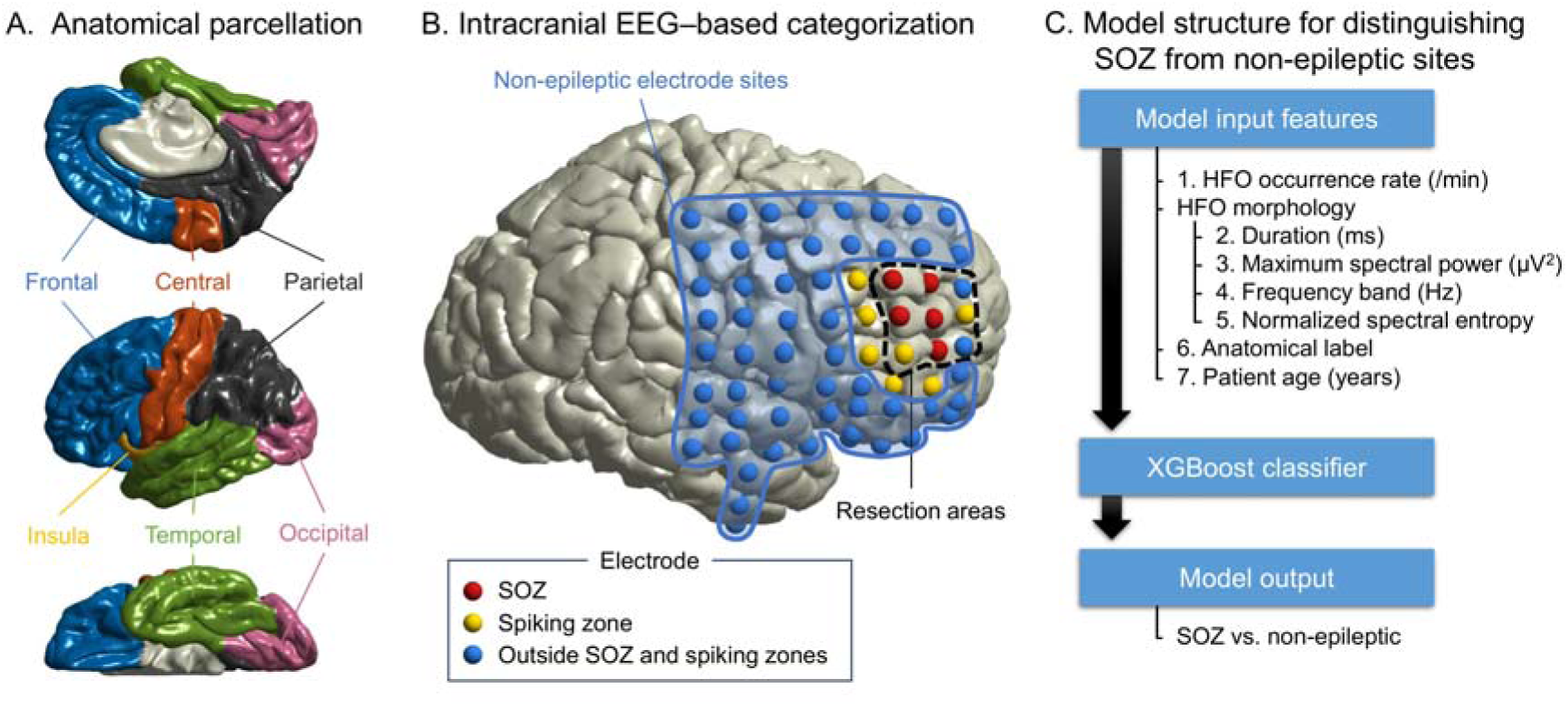
Schematic illustration defining key terms used in this study. **A.** *Anatomical parcellation*: Each electrode site was assigned an anatomical label based on its location on the cortical surface. **B.** *Intracranial EEG–based categorization*: Non-epileptic electrode sites were defined as those located outside the seizure onset zone (SOZ), spiking zones, and resection areas^23^. **C.** *Model structure for distinguishing SOZ from non-epileptic sites*: Seven features were used as inputs to the XGBoost classifier.

### Resection and postoperative outcome

The extent of resection was determined by a multidisciplinary epilepsy surgery team at each institution. In general, the surgical team aimed to remove the SOZ and the adjacent MRI-visible lesion, if present. When habitual seizures were not captured, the resection included the MRI-visible lesion and cortical regions that frequently generated interictal spike discharges. If eloquent cortex was identified within the planned resection, the final margin was determined after discussion with the patient or their guardian regarding the risks and benefits of resection versus preservation. Intraoperative photographs obtained upon completion of resection were used to verify that the intended resection was achieved. The proportion of the resected cortical surface relative to the total hemispheric surface area was quantified using the FreeSurfer program^36^. In our prior analysis of 89 patients, the spatial extents of resection estimated from intraoperative photographs and postoperative MRI were highly correlated^36^ (ρ = 0.95, p < 0.001). Postoperative seizure outcome was classified according to the International League Against Epilepsy (ILAE) scale as Class 1 or Class ≥2.

### Training and internal cross-validation of the SOZ classification model

Using training data from derivation-cohort patients who developed a habitual seizure during extraoperative iEEG recording and achieved an ILAE Class 1 outcome, we trained the model to classify SOZ versus non-epileptic electrode sites using the XGBoost algorithm, implemented in Python 3.12.3 (xgboost library version 2.1.3; https://xgboost.ai/). Postoperative seizure control supports the validity of the SOZ as a proxy for the epileptogenic zone in this cohort. Non-epileptic electrode sites were defined as those located outside the SOZ, spiking zone, and resection areas^23^ (**Figure 1B**). The comprehensive HFA-based model for SOZ classification included seven features: HFA occurrence rate; HFA morphological characteristics (duration, maximum spectral power, frequency at maximum spectral power, and normalized spectral entropy); electrode location; and patient age (**Figure 1C**). The model assigned an SOZ probability score (range: 0 to 1) to each electrode site. To mitigate overfitting, we employed nested 10-fold cross-validation for hyperparameter tuning and model evaluation, as detailed in **Table S1**. To evaluate how accurately the SOZ probability score identified clinician-defined SOZ sites, we computed the Area Under the Receiver Operating Characteristic Curve (AUROC) and the Area Under the Precision–Recall Curve (AUPRC). AUPRC is informative when class imbalance exists between SOZ and non-epileptic electrode sites^43^.

To exclude the possibility that SOZ classification was driven by spurious patterns captured by the XGBoost algorithm, we evaluated model performance using pseudo-SOZ labels. These pseudo-SOZ labels were randomly assigned to 16.8% of electrode sites, matching the proportion of clinician-defined SOZ sites in the derivation cohort for the SOZ classification. We constructed 100 models to classify pseudo-SOZ sites and used a permutation test to compare their AUROC performance with that of models trained on clinician-defined SOZ labels.

To evaluate the contribution of each feature to the classification of clinician-defined SOZ sites, we applied SHapley Additive exPlanations (SHAP), which provide a unified framework for estimating feature importance based on cooperative game theory^44^. Both global and feature-specific contributions were visualized using SHAP summary and individual plots.

We also assessed how accurately the HFA occurrence rate alone identified clinician-defined SOZ sites by computing the AUROC and AUPRC. The DeLong test and a bootstrap paired t-test were applied to AUROC and AUPRC, respectively, to determine whether the SOZ probability score derived from the comprehensive HFA-based model classified clinician-defined SOZ sites more accurately than the HFA occurrence rate alone.

### External validation of the SOZ classification model

Using the aforementioned comprehensive HFA-based model derived from the derivation cohort, we computed the SOZ probability score for each electrode site in the temporal and geographical external validation cohorts. We then computed the AUROC and AUPRC to evaluate how accurately the model-derived SOZ probability score—and, for comparison, the HFA occurrence rate alone—identified clinician-defined SOZ sites in these independent cohorts.

### Internal cross-validation of the outcome classification model

We evaluated whether more inclusive resection of regions exhibiting excessive abnormalities—reflected by either high SOZ probability scores or elevated HFA occurrence rates—was associated with achieving ILAE Class 1 outcomes in the derivation cohort. The extent to which regions with abnormal biomarker values were resected was quantified using a summary measure, termed the ‘biomarker difference,’ defined as the difference between the mean biomarker value across resected electrode sites and that across preserved sites, as described in our previous study^34^. A higher biomarker difference indicates that regions with markedly elevated biomarker values were preferentially resected, whereas regions with relatively lower values were preserved. Using AUROC and AUPRC, we evaluated how accurately the ‘SOZ probability biomarker difference’—and, separately, the ‘HFA rate biomarker difference’—classified patients who achieved an ILAE Class 1 outcome.

Subsequently, we evaluated whether adding the biomarker difference as a predictor improved the accuracy of a logistic regression model based on information available in standard care for classifying patients who achieved an ILAE Class 1 outcome. The standard-care logistic regression model incorporated the following 10 predictor variables: age, sex, presence of daily seizures, number of oral ASMs, affected hemisphere, presence of an MRI lesion, whether iEEG recording captured habitual seizures, whether the clinician-defined SOZ was incompletely removed, the necessity of extratemporal lobe resection, and the extent of resection^34^. To mitigate overfitting, we performed a leave-one–patient-out analysis. Model performance—with and without the biomarker difference—was quantified using the AUROC and AUPRC.

### External validation of the outcome prediction model

Using SOZ probability scores derived from the comprehensive HFA-based model, we computed the biomarker difference for each patient in the temporal and geographical external validation cohorts. We then calculated the AUROC and AUPRC to evaluate how accurately the ‘SOZ probability biomarker difference’—and, for comparison, the ‘HFA rate biomarker difference’—predicted patients with an ILAE Class 1 outcome in these independent external validation cohorts.

### Exploring alternative summary measures of resection completeness

Because no universally accepted summary metric exists to quantify the overall extent of resection of regions exhibiting abnormally elevated biomarker values, we evaluated outcome-classification performance using alternative summary measures beyond the ‘biomarker difference’^34^. Specifically, we assessed the predictive utility of models incorporating each of the following metrics: ‘difference index’^17^, ‘resection ratio’^45^, ‘critical resection percentage’^46^, and ‘distinguishability statistic’^47^. The definitions of these measures are provided in **Figure S2**.

### Examining the MRI-based etiology linked to impaired model performance

We evaluated whether model performance was attributable to specific etiologies. To this end, using all data from the external validation cohorts, we computed the AUROC for classifying clinician-defined SOZ and for predicting ILAE Class 1 outcome within four MRI-based etiological subgroups: non-lesional epilepsy, tumor, dysplasia (excluding encephalomalacia), and encephalomalacia.

### Statistical analyses

Continuous variables were summarized as mean ± standard deviation (SD) or as median with interquartile range (IQR), as appropriate. Differences in median HFA occurrence rate and morphological features between SOZ and non-epileptic electrode sites were assessed using the Wilcoxon rank-sum test. Age-related changes in these differences were evaluated using Spearman’s rho. AUROC was compared using the DeLong test, and AUPRC using the bootstrap procedure. A two-sided significance threshold of p < 0.0125 was applied using a Bonferroni correction for four comparisons across the HFA detectors (STE, SLL, HIL, and MNI). The Wilcoxon rank-sum test, Spearman’s rho, logistic regression analysis, and DeLong test were performed using SPSS v30 (IBM, Armonk, NY). Bootstrap procedures and permutation tests were conducted using MATLAB R2023b (MathWorks, Natick, MA, USA).

## Results

### Patient Characteristics

At the Detroit Medical Center, 142 patients (15,464 electrodes) satisfied the eligibility criteria for the derivation cohort (**Table S2**). Among these 142 patients, 79 experienced habitual seizure events during extraoperative iEEG monitoring and achieved an ILAE Class 1 outcome following resective surgery; in these 79 patients, 4,905 electrode sites classified as either SOZ (822 electrode sites) or non-epileptic (4,083 electrode sites) were used for training the SOZ classification model. At the same center, 35 patients (3,934 electrodes) satisfied the eligibility criteria for the temporal external validation cohort. At Tohoku University and NCNP, 26 patients (1,644 electrodes) and 30 patients (1,897 electrodes), respectively, satisfied the eligibility criteria for the geographical external validation cohorts. **Figure S3** shows the spatial distribution of electrode sites in each cohort. A total of 5,142,891 HFA events were identified across artifact-free electrodes from all cohorts (STE detector: 403,314; SLL: 2,327,849; HIL: 1,999,046; MNI: 412,682).

## Difference in HFA features between SOZ and non-epileptic electrode sites

Figure 2 illustrates the distribution of HFA features in the training dataset comprising 79 patients. Differences between SOZ and non-epileptic sites, assessed using the Wilcoxon signed-rank test (**Table 1**), are summarized below.

- Occurrence rate: Across all four detectors, the median HFA occurrence rate was higher in SOZ sites, with large effect sizes ranging from +0.72 to +0.77.
- Normalized spectral entropy: The median entropy of STE-, HIL-, and MNI-defined HFA was higher in SOZ sites (effect sizes: +0.47 to +0.68), whereas the median entropy of SLL-defined HFA was lower in SOZ sites (effect size: −0.38).
- Maximum spectral power: The median spectral power of STE-, SLL-, and HIL-defined HFA was higher in SOZ sites (effect sizes: +0.44 to +0.68), whereas MNI-defined HFA showed no significant difference between SOZ and non-epileptic sites (effect size: +0.18).
- Frequency band: The median frequency band of STE- and HIL-defined HFA was higher in SOZ sites (effect sizes: +0.18 to +0.47), whereas SLL- and MNI-defined HFA exhibited no significant difference (effect sizes: −0.04 to −0.23).
- Duration: The median duration of SLL-defined HFA was higher in SOZ sites (effect size: +0.74), whereas HFA defined by the other detectors showed no significant difference (effect sizes: +0.06 to +0.27).

**Figure 2.**
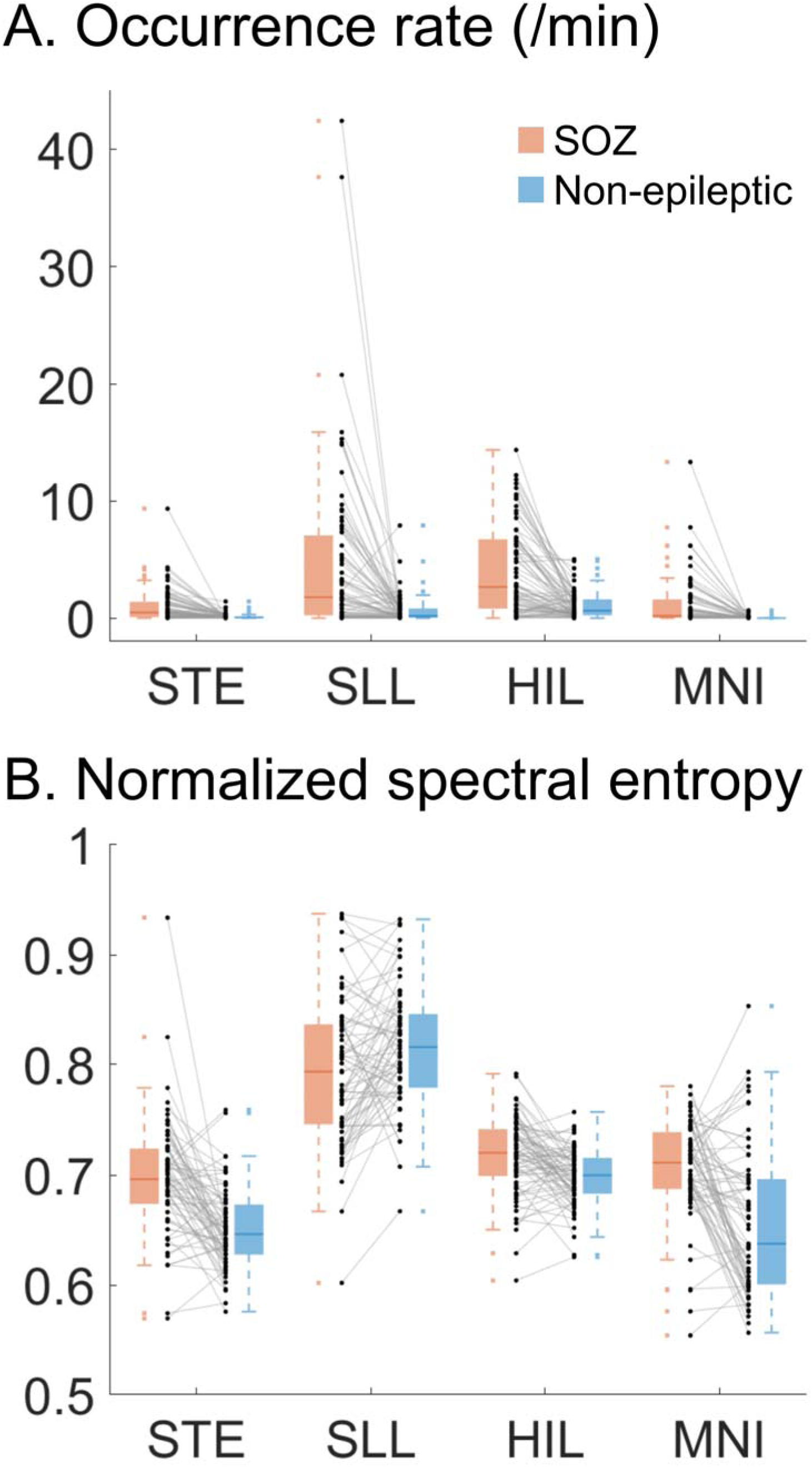
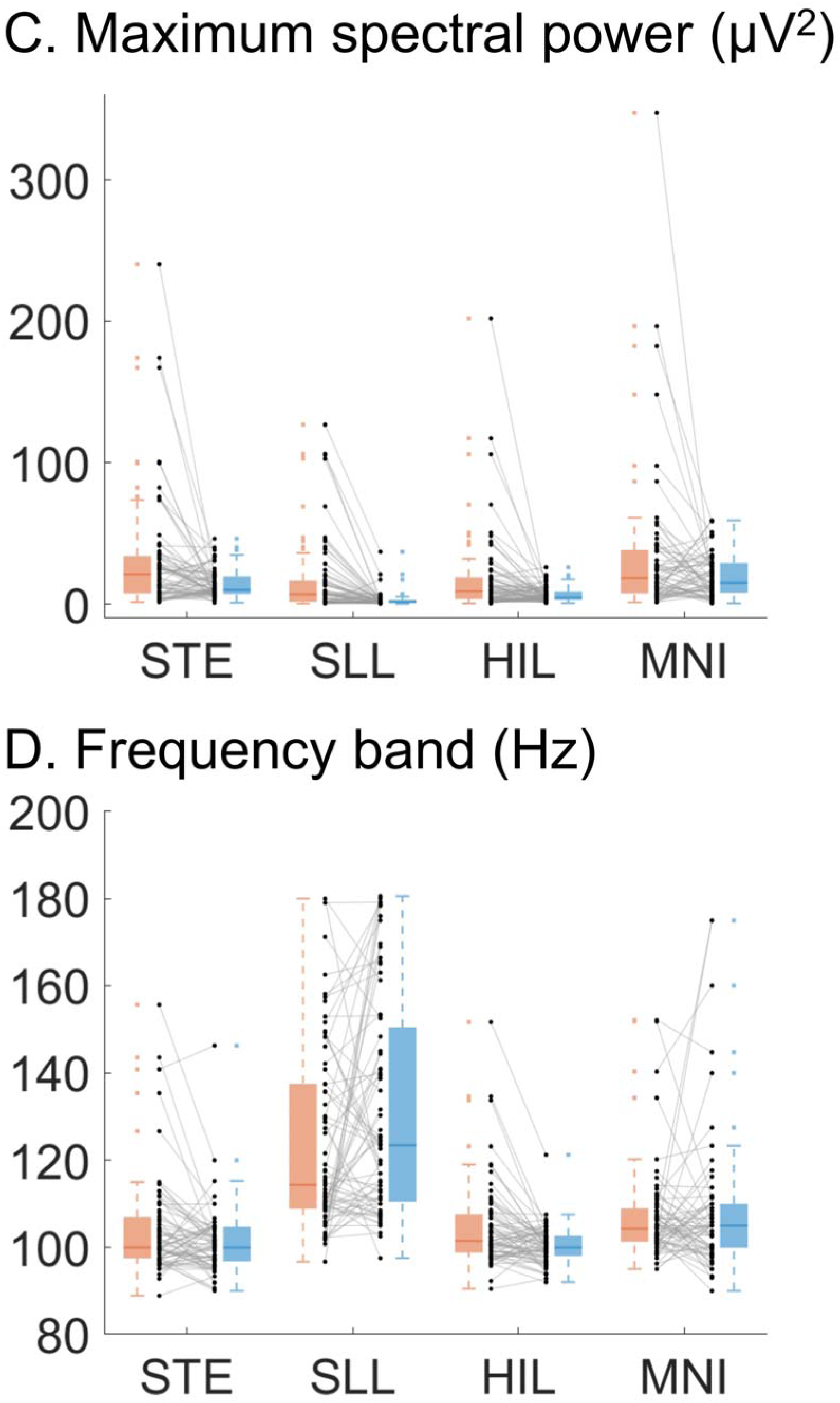
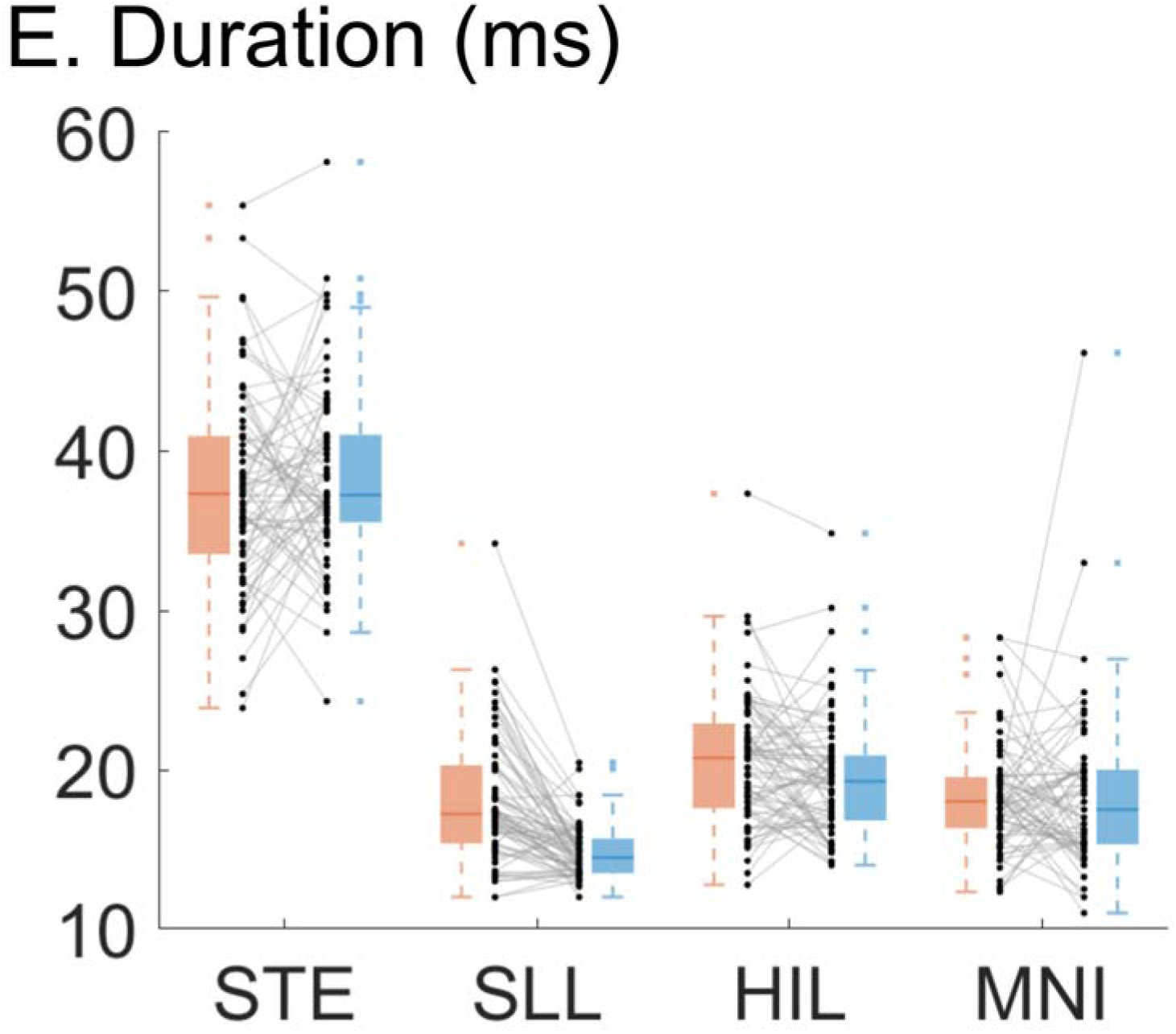
High-frequency activity (HFA) features at seizure onset and non-epileptic sites. Paired dot plots and corresponding box plots show differences of HFA features between seizure onset zone (SOZ) and non-epileptic sites. Black dots represent medians computed per patient among the 79 patients in the derivation cohort, and box plots show median and interquartile range of black dots at SOZ (red) and non-epileptic sites (blue). The broken lines extending from the box (whiskers) indicate the range of the data within 1.5 times from the interquartile range. **A.** Occurrence rate. **B.** Normalized spectral entropy. **C.** Maximum spectral power. **D.** Frequency band. **E.** Duration. STE detector: Short-Time Energy detector. SLL: Short Line Length. HIL: Hilbert. MNI: Montreal Neurological Institute.

**Table 1.** High-frequency activity (HFA) features at seizure onset and non-epileptic sites. A. Occurrence rate (medians computed per patient) at seizure onset zone and non-epileptic sites among the 79 patients in the derivation cohort. **B.** Normalized spectral entropy. **C.** Maximum spectral power. **D.** Frequency band. **E.** Duration. STE detector: Short-Time Energy detector. SLL: Short Line Length. HIL: Hilbert. MNI: Montreal Neurological Institute. The effect size (r), calculated as Z/√N, and the p-value were derived from the Wilcoxon signed-rank test. Here, Z denotes the standardized test statistic, and N repr esents the number of patients contributing SOZ–non-epileptic pairs. Effect size thresholds were interpreted as small (r≈0.1), medium (r≈0.3), and large (r≥0.5).

|  | Median (IQR) |  | Effect size<br>(r) | p-value |
| --- | --- | --- | --- | --- |
|  | SOZ | Non epileptic |  |  |
| Occurrence rate (/min) |  |  |  |  |
| STE | 0.47<br>(0.09–1.40) | 0.05<br>(0.02–0.13) | 0.74 | <0.001 |
| SLL | 1.78<br>(0.24–7.06) | 0.19<br>(0.04–0.82) | 0.75 | <0.001 |
| HIL | 2.65<br>(0.81–6.74) | 0.63<br>(0.24–1.6) | 0.72 | <0.001 |
| MNI | 0.20<br>(0.03–1.59) | 0.00<br>(0.00–0.01) | 0.77 | <0.001 |
| Normalized spectral entropy |  |  |  |  |
| STE | 0.70<br>(0.67–0.72) | 0.65<br>(0.63–0.67) | 0.68 | <0.001 |
| SLL | 0.79<br>(0.75–0.84) | 0.82<br>(0.78–0.85) | -0.38 | 0.001 |
| HIL | 0.72<br>(0.70–0.74) | 0.70<br>(0.68–0.72) | 0.47 | <0.001 |
| MNI | 0.71<br>(0.69–0.74) | 0.64<br>(0.60–0.70) | 0.56 | <0.001 |
| Maximum spectral power (μV<sup>2</sup>) |  |  |  |  |
| STE | 21.06<br>(7.57–34.13) | 10.19<br>(7.09–19.60) | 0.44 | <0.001 |
| SLL | 6.92<br>(1.55–16.57) | 1.41<br>(0.80–3.09) | 0.68 | <0.001 |
| HIL | 9.11<br>(3.63–18.83) | 4.71<br>(2.98–8.93) | 0.47 | <0.001 |
| MNI | 18.4<br>(7.52–38.39) | 15.13<br>(7.95–29.11) | 0.18 | 0.132 |
| Frequency band (Hz) |  |  |  |  |
| STE | 100.00<br>(97.44–106.91) | 100.00<br>(96.73–104.72) | 0.18 | 0.011 |
| SLL | 114.38<br>(108.88–137.55) | 123.44<br>(110.48–150.42) | -0.23 | 0.05 |
| HIL | 101.45<br>(98.80–107.56) | 100.00<br>(98.00–102.69) | 0.47 | <0.001 |
| MNI | 104.29<br>(101.25–108.99) | 105.00<br>(99.96–110.00) | -0.04 | 0.759 |
| Duration (ms) |  |  |  |  |
| STE | 37.28<br>(33.55–40.91) | 37.20<br>(35.49–40.99) | 0.12 | 0.322 |
| SLL | 17.23<br>(15.38–20.28) | 14.48<br>(13.50–15.68) | 0.74 | <0.001 |
| HIL | 20.74<br>(17.60–22.90) | 19.28<br>(16.82–20.90) | 0.27 | 0.023 |
| MNI | 18.02<br>(16.34–19.55) | 17.50<br>(15.31–20.00) | 0.06 | 0.646 |

Figure S4 shows the median HFA feature values at SOZ and non-epileptic sites across patient age. The magnitude of the differences in HFA occurrence rate and frequency band between SOZ and non-epileptic sites was greater in younger patients, with the pattern varying by HFA detector. For example, younger age was associated with a larger difference in SLL-defined HFA occurrence rate between SOZ and non-epileptic sites (Spearman’s rho = −0.39, p = 0.001). Similarly, younger age was associated with larger differences in STE- and MNI-defined HFA frequency bands between SOZ and non-epileptic sites (STE: rho = −0.34, p = 0.003; MNI: rho = −0.32, p = 0.009). No other HFA features demonstrated significant age-dependent differences between SOZ and non-epileptic sites.

### SOZ classification performance

The model-derived SOZ probability score differentiated clinician-defined SOZ from non-epileptic sites with AUROC values ranging from 0.79 to 0.85 in internal nested cross-validation, 0.77 to 0.86 in the temporal external validation cohort, and 0.71 to 0.75 in the geographical external validation cohorts. AUROC values varied across HFA detectors. Across the internal cross-validation, temporal external validation, and NCNP geographical external validation cohorts, the SOZ probability score showed improved classification of clinician-defined SOZ compared with HFA occurrence rate alone (DeLong test, p < 0.0125; Figure 3). This improvement was evident for HIL-, SLL-, and STE-defined HFA, with AUROC increases of up to 12.7%. In contrast, the incremental benefit of the SOZ probability score over HFA occurrence rate alone was modest for MNI-defined HFA, with AUROC increases of 5.8% or less. The SOZ probability score and HFA occurrence rate alone showed comparable SOZ classification performance in the Tohoku geographical external validation cohort.

**Figure 3.**
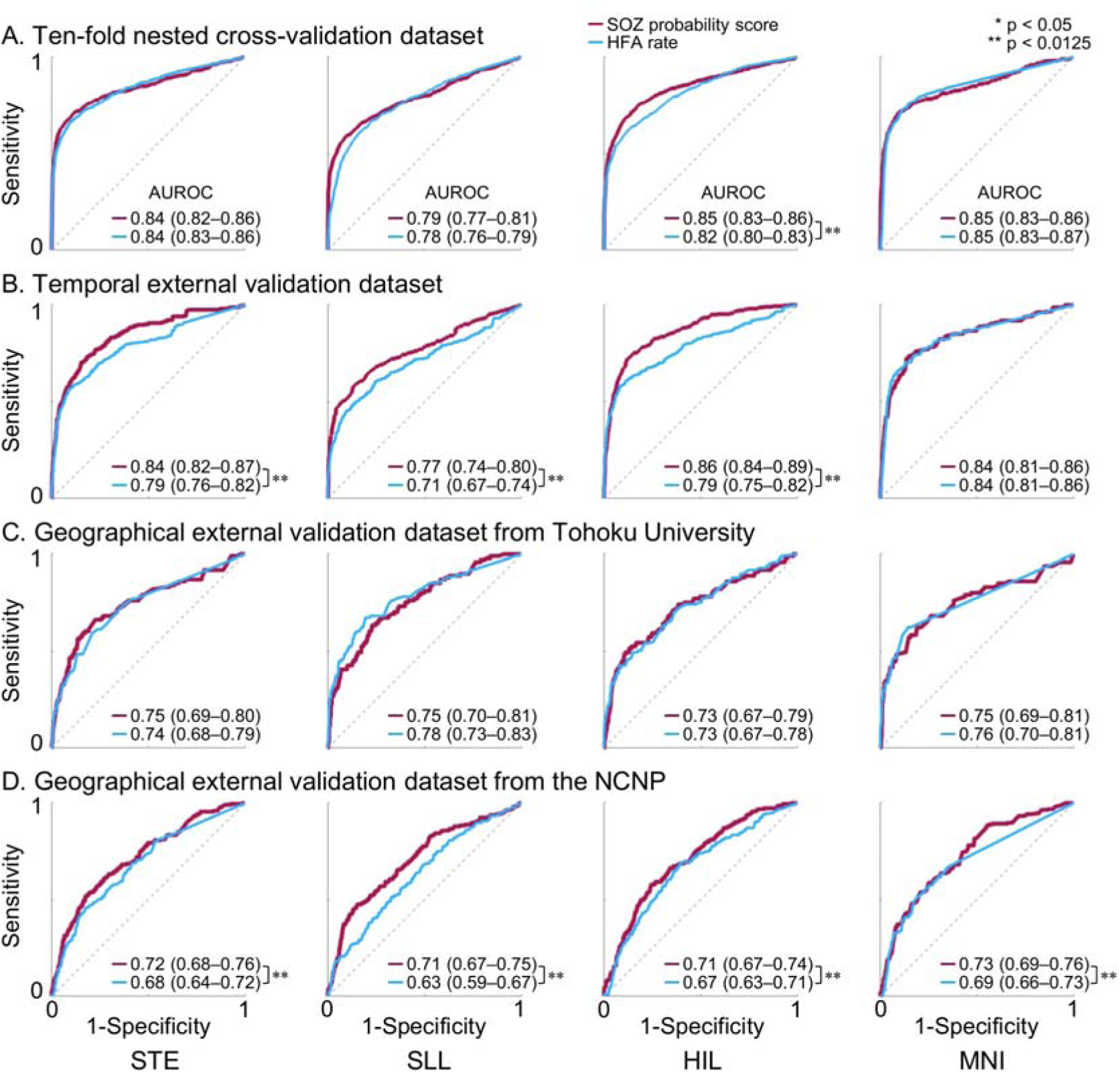
Accuracy of intracranial EEG biomarker–based classification of the seizure onset zone (SOZ). Classification performance for distinguishing SOZ from non-epileptic sites using the model-derived SOZ probability score (red) and the high-frequency activity (HFA) occurrence rate alone (blue), quantified by the area under the receiver operating characteristic curve (AUROC). **A.** Ten-fold nested cross-validation within the internal cohort. **B.** Temporal external validation dataset. **C.** Geographical external validation dataset from Tohoku University. **D.** Geographical external validation dataset from the National Center of Neurology and Psychiatry (NCNP). STE detector: Short-Time Energy detector. SLL: Short Line Length. HIL: Hilbert. MNI: Montreal Neurological Institute. * p < 0.05; ** p < 0.0125 derived from the DeLong test.

Figure S5 shows the AUPRC for classification of clinician-defined SOZ versus non-epileptic sites. Across the internal cross-validation, temporal external validation, and NCNP geographical external validation cohorts, the SOZ probability score achieved a higher AUPRC than HFA occurrence rate alone (DeLong test, p < 0.0125), although the magnitude of improvement varied across HFA detectors. The maximum AUPRC gain provided by the SOZ probability score reached 30.8%. In contrast, in the Tohoku geographical external validation cohort, the SOZ probability score did not outperform HFA occurrence rate alone in terms of AUPRC.

### Confirmation of model robustness using pseudo-SOZ labels

The nested cross-validation AUROC for classifying pseudo-SOZ sites was 0.50 (**Table S3**), whereas that for clinician-defined SOZ sites ranged from 0.79 to 0.85, depending on the HFA detector. The difference in AUROC between clinician-defined SOZ and pseudo-SOZ labels was statistically significant for all detectors tested (permutation test, p = 0.0099). This marked contrast indicates that the model’s accurate classification of clinician-defined SOZ sites cannot be attributed to spurious patterns learned by the XGBoost algorithm.

#### Features driving SOZ classification

The SHAP summary plots (Figure 4A) indicate that the HFA occurrence rate was among the most influential features for classification of clinician-defined SOZ sites using the SOZ probability score derived from XGBoost. Higher occurrence rates were associated with higher SHAP values, reflecting a stronger contribution to classifying SOZ sites (Figure 4B). Normalized spectral entropy also demonstrated substantial influence: entropy values in the mid-range (∼0.7–0.8) yielded elevated SHAP values, whereas both lower and higher entropy values contributed less (Figure 4C). Maximum spectral power was an influential feature as well, particularly for SLL-defined HFA, with higher power corresponding to higher SHAP values (Figure 4D). In contrast, the HFA frequency band and event duration were among the least influential features for SOZ classification (Figure 4A). Younger patients tend to have higher SHAP value (Figure 4G), reflecting that younger patients had a higher proportion of SOZ sites.

**Figure 4.**
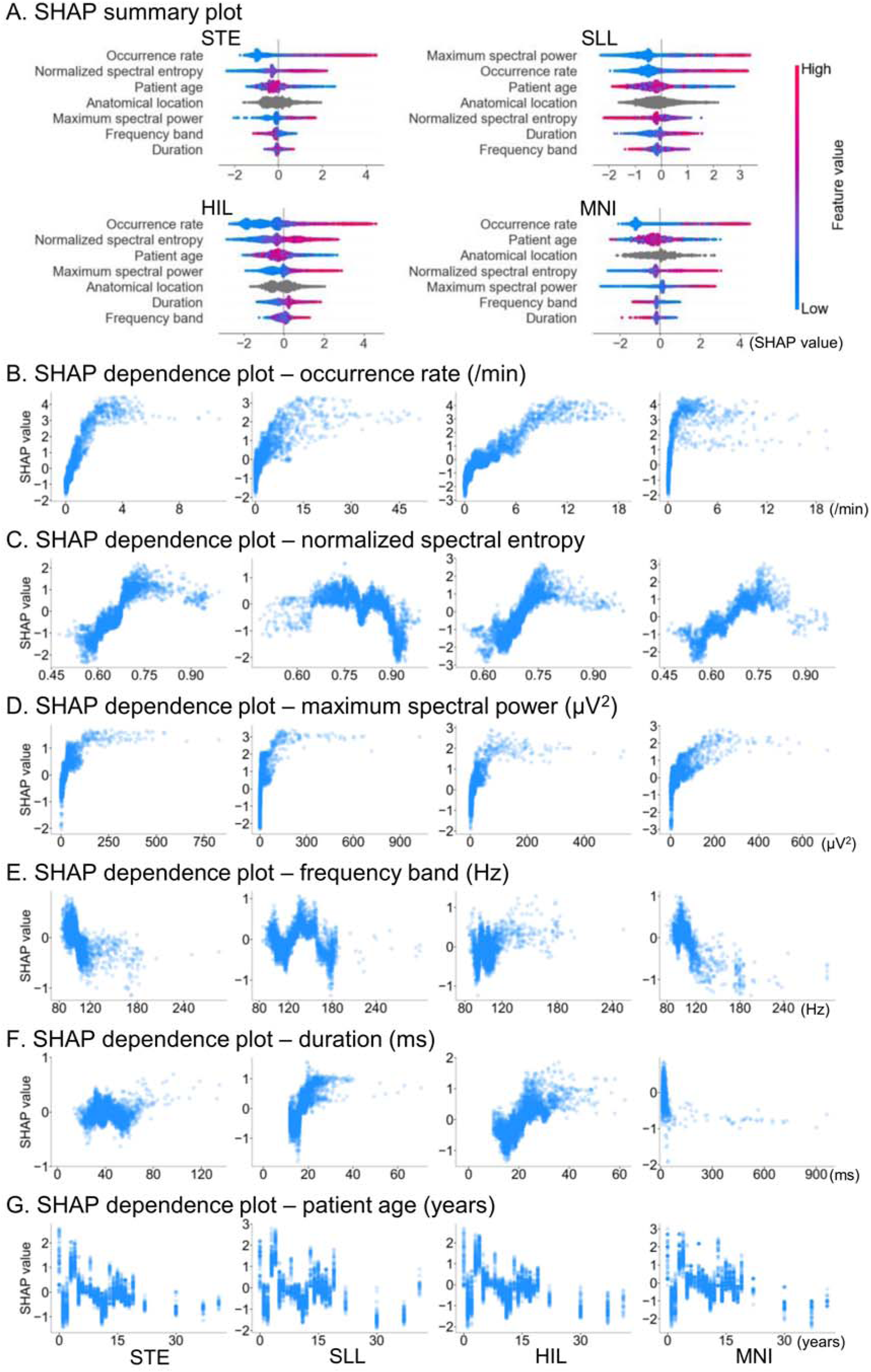
Features driving SOZ classification. **A.** SHapley Additive exPlanations (SHAP) summary plot illustrating the contribution of each feature to XGBoost classification of SOZ versus non-epileptic sites in the 79 model-development patients. Features are ranked from top (most influential) to bottom (least influential) by mean absolute SHAP values. The color bar shows feature values (red = higher, blue = lower). Positive SHAP values indicate contributions toward SOZ classification, whereas negative values indicate contributions toward non-epileptic classification. SHAP dependence plots illustrate how feature values relate to their corresponding SHAP values for individual data points. **B.** Occurrence rate. **C.** Normalized spectral entropy. **D.** Maximum spectral power. **E.** Frequency band. **F.** Duration. **G.** Patient age. STE detector: Short-Time Energy detector. SLL: Short Line Length. HIL: Hilbert. MNI: Montreal Neurological Institute.

### Internal validation of seizure outcome classification models

Univariate logistic regression in the 142-patient derivation cohort showed that greater resection completeness of regions with elevated SOZ probability scores—indexed by the enhanced ‘biomarker difference’—was associated with a higher likelihood of achieving an ILAE class 1 outcome (p ≤ 0.004; odds ratio [OR] ranging from 9.8 to 36.9 depending on the HFA detector used; **Table S4**). In multivariate logistic regression, the enhanced ‘SOZ probability biomarker difference’ remained independently associated with ILAE class 1 outcome, after accounting for 10 predictor variables routinely available in standard clinical management (p ≤ 0.01; OR: 11.3 to 87.5; **Table S5**). In contrast, neither the univariate nor the multivariate logistic regression model indicated that the ‘HFA rate biomarker difference’ was associated with achieving an ILAE class 1 outcome (p ≥0.013; **Tables S6–S7**).

In the same derivation cohort, the ‘SOZ probability biomarker difference’ classified patients achieving an ILAE class 1 outcome with an AUROC ranging from 0.65 to 0.70, whereas the ‘HFA rate biomarker difference’ yielded an AROUC ranging from 0.53 to 0.59 (Figure 5A). The ‘SOZ probability biomarker difference’ achieved a significantly higher AUROC than the ‘HFA rate biomarker difference’ when STE-, SLL-, and HIL-defined HFA events were analyzed (p < 0.0125).

**Figure 5.**
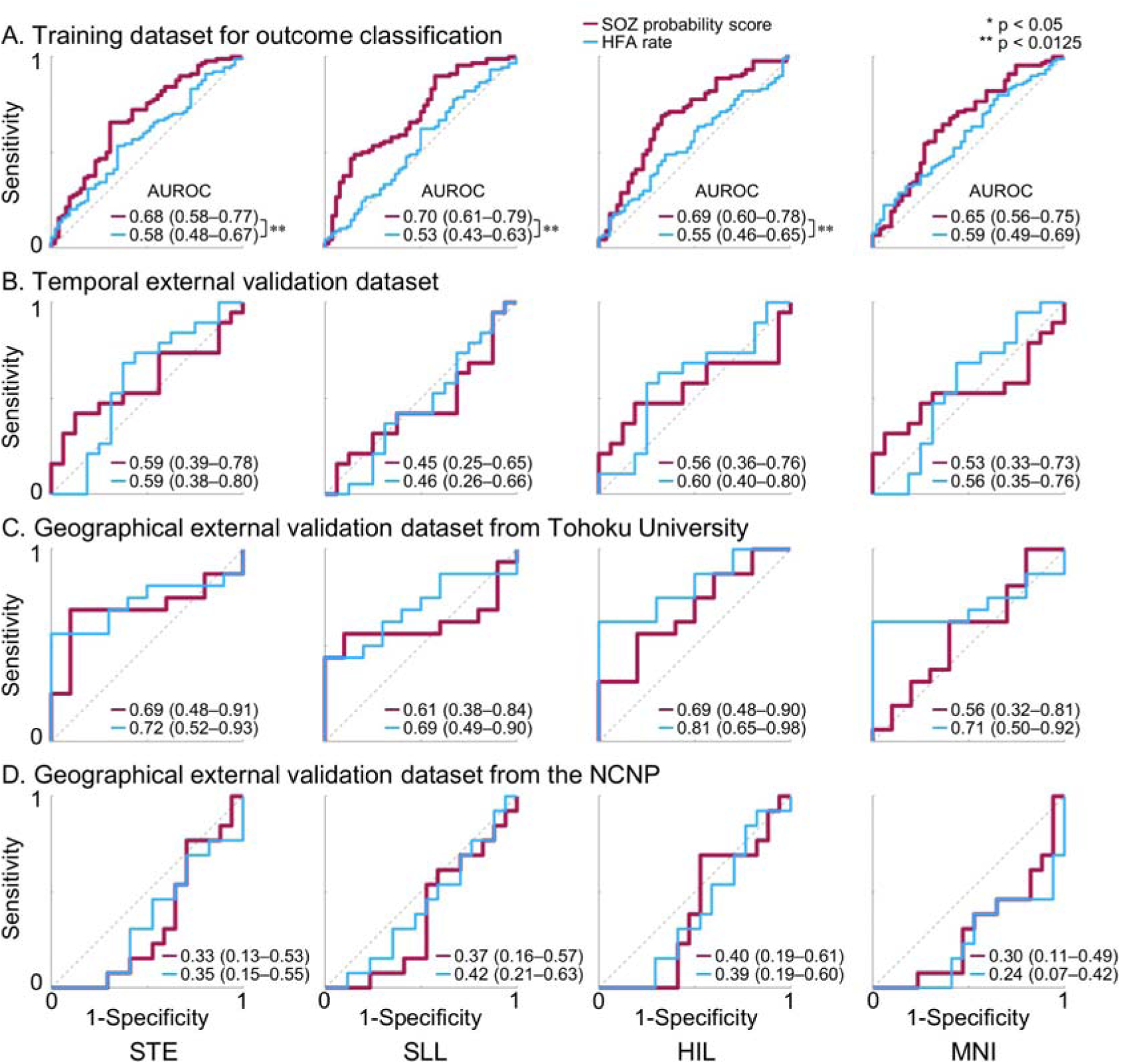
Accuracy of intracranial EEG biomarkers for classification of seizure outcomes. Receiver operating characteristic (ROC) curves illustrate the performance of classifying patients who achieved an ILAE class 1 outcome using the ‘seizure onset zone (SOZ) probability biomarker difference’ (red) and the ‘high-frequency activity (HFA) rate biomarker difference’ (blue). For each curve, the area under the ROC curve (AUROC) with its 95% confidence interval (95% CI) is shown. The area under the precision–recall curve (AUPRC) is provided in **Figure S6**. STE detector: Short-Time Energy detector. SLL: Short Line Length. HIL: Hilbert. MNI: Montreal Neurological Institute. * p < 0.05; ** p < 0.0125 derived from the DeLong test.

Another ROC analysis using leave-one-out internal cross-validation showed that incorporating the ‘SOZ probability biomarker difference’ into the standard-care logistic regression model improved seizure outcome classification (Figure 6B). The standard-care model alone classified ILAE class 1 outcome with an AUROC of 0.58, whereas adding the ‘SOZ probability biomarker difference’ increased the AUROC to 0.63–0.70 across detectors (Figure 6B). The magnitude of this improvement was significant for the STE, SLL, and HIL detectors (p < 0.0125).

**Figure 6.**
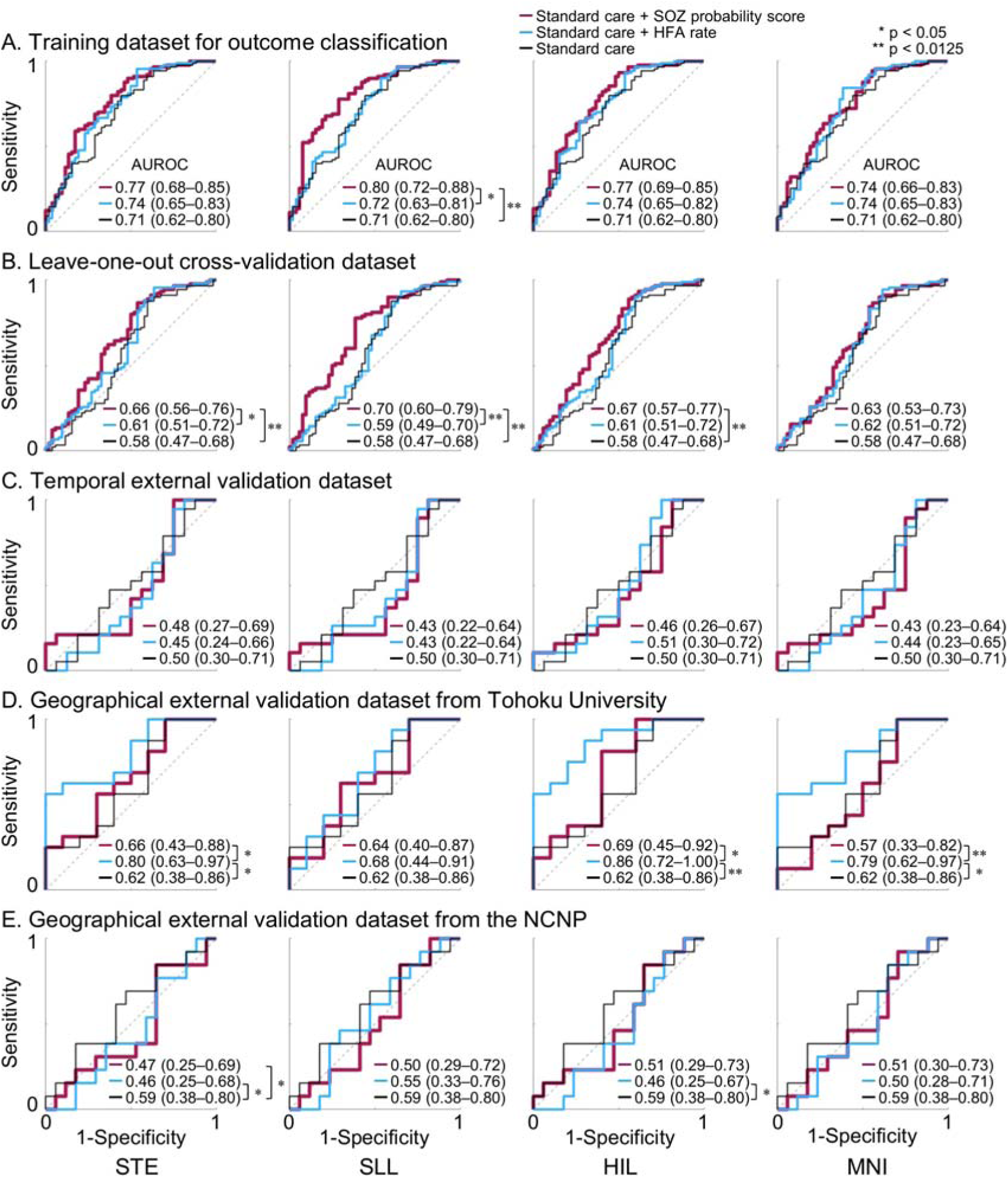
Additive value of intracranial EEG biomarkers for classification of seizure outcomes. Receiver operating characteristic (ROC) curves illustrate the performance of three logistic regression models for classifying patients who achieved an ILAE class 1 outcome. The red curve represents the model incorporating the ‘seizure onset zone (SOZ) probability biomarker difference’ in addition to the standard-care model. The blue curve represents the model incorporating the ‘high-frequency activity (HFA) rate biomarker difference’ in addition to the standard-care model. The black curve represents the standard-care model alone. For each curve, the area under the ROC curve (AUROC) with its 95% confidence interval (95% CI) is shown. The corresponding area under the precision–recall curve (AUPRC) is provided in **Figure S7**. STE detector: Short-Time Energy detector. SLL: Short Line Length. HIL: Hilbert. MNI: Montreal Neurological Institute. * p < 0.05; ** p < 0.0125 derived from the DeLong test.

### External validation of seizure outcome classification models

The model-based SOZ probability did not accurately predict, nor did it improve the prediction of, ILAE class 1 outcomes in either the temporal or geographical external cohorts. The ‘SOZ probability biomarker difference’ yielded AUROC values ranging from 0.30 to 0.69, depending on the cohorts analyzed and HFA detectors used (Figures 5B**–5D**). In comparison, the logistic regression model using standard-care variables alone had an AUROC of 0.50–0.62; after adding the ‘SOZ probability biomarker difference’, the AUROC ranged from 0.43 to 0.69 (Figures 6C**–6E**).

Notably, in one geographical cohort (NCNP), the ‘SOZ probability biomarker difference’ was inversely related to outcome. When MNI-defined HFA events were analyzed, the AUROC was 0.30 (95% CI: 0.11–0.49), indicating that greater resection of areas with higher SOZ probability was associated with a lower likelihood of achieving ILAE class 1 outcome in this cohort (Figure 5D). Likewise, when STE-defined HFA events were analyzed, incorporating the ‘SOZ probability biomarker difference’ reduced the AUROC by the standard-care model from 0.59 to 0.47 (p = 0.019) in this cohort (Figure 6E).

### Model performance across MRI-defined etiologies

The external validation cohorts included 26 patients with non-lesional epilepsy, 13 with tumor, 28 with dysplasia (excluding encephalomalacia), and 10 with encephalomalacia. Both the model-based SOZ probability score and HFA occurrence rate demonstrated SOZ classification performance above chance (p < 0.0125; Figure 7A). The SOZ probability score classified clinician-defined SOZ sites with AUROC values ranging from 0.68 to 0.86, whereas the HFA rate yielded AUROC values ranging from 0.64 to 0.81, depending on the HFA detector used.

**Figure 7.**
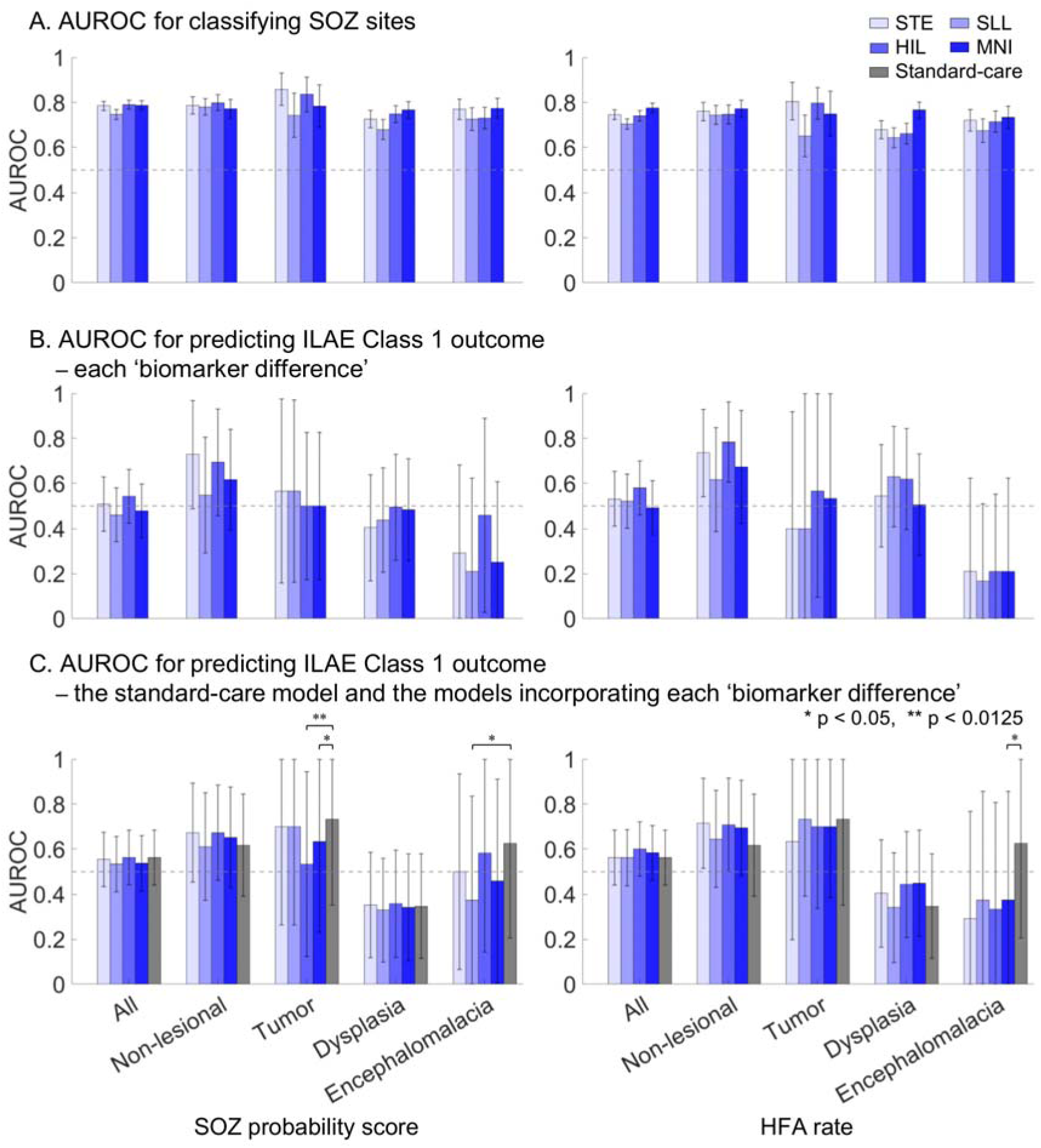
Performance of seizure onset zone classification and postoperative outcome prediction across MRI-defined etiologies in the external validation cohorts (n = 91). **A.** Area under the receiver operating characteristic curve (AUROC) for classifying seizure onset zone (SOZ) sites within each MRI-defined etiology. Left: classification using the SOZ probability score derived from comprehensive high-frequency activity (HFA) model. Right: classification using HFA rate alone. **B.** AUROC for classifying patients with ILAE Class 1 outcome. Left: classification using the ‘SOZ probability biomarker difference.’ Right: classification using the ‘HFA rate biomarker difference.’ **C.** AUROC for predicting ILAE Class 1 outcome using the standard-care model and the models incorporating each ‘biomarker difference.’ The horizontal dotted line represents the AUROC reference value (0.5), and the error bars indicate the 95% confidence interval. STE detector: Short-Time Energy detector. SLL: Short Line Length. HIL: Hilbert. MNI: Montreal Neurological Institute. * p < 0.05; ** p < 0.0125, as determined by the DeLong test in comparison with the standard-care model.

In contrast, SOZ probability score–based prediction of postoperative seizure outcome varied substantially across MRI-defined etiologies. Performance was highest in patients with non-lesional epilepsy and lowest in those with encephalomalacia. Specifically, the ‘SOZ probability biomarker difference’ classified ILAE Class 1 outcomes with AUROC values of 0.55–0.73 in non-lesional epilepsy, 0.50–0.57 in tumor, 0.40–0.49 in dysplasia, and 0.21–0.46 in encephalomalacia, depending on the HFA detector used (Figure 7B). The ‘HFA rate biomarker difference’ showed a similar pattern, yielding the highest AUROC values in non-lesional epilepsy (0.62–0.78) and the lowest in encephalomalacia (0.17–0.21). In encephalomalacia, greater preferential resection of regions with higher SOZ probability or elevated HFA rate was associated with lower likelihood of achieving ILAE Class 1 outcome.

Adding the ‘SOZ probability biomarker difference’ to the standard-care model shifted AUROC for predicting ILAE Class 1 outcome from 0.62 to 0.60–0.67 in non-lesional epilepsy (Figure 7C). Similarly, adding the ‘HFA rate biomarker difference’ increased AUROC from 0.62 to 0.65–0.72 in non-lesional epilepsy. In contrast, incorporation of either biomarker difference reduced AUROC from 0.62 to 0.29–0.58 in patients with encephalomalacia.

### Outcome-prediction performance across summary measures of resection completeness

Figures S8–S11 summarize ROC analyses based on the other summary measures of resection completeness—including the ‘difference index’^17^, ‘resection ratio’^45^, ‘critical resection percentage’^46^, and ‘distinguishability statistic’^47^. These measures yielded outcome-prediction performance comparable to that of the ‘biomarker difference’ metric^34^.

## Discussion

### Summary

The current study demonstrates that interictal HFA encodes robust information about the spatial extent of habitual seizure onset. By integrating HFA occurrence rates with morphological features from training iEEG data obtained from patients who achieved postoperative seizure freedom, we derived an SOZ probability score that reliably classified clinician-defined SOZ sites. The accurate classification of SOZ sites across both temporal and geographical external cohorts indicates that this model-derived score captures neurophysiological signatures of the epileptogenic zone. However, the SOZ probability score did not translate into consistent prediction of postoperative seizure outcomes across external cohorts. Notably, prognostic performance varied by etiology: in MRI-nonlesional epilepsy, both HFA rate and SOZ probability score showed moderate predictive value, whereas in encephalomalacia—characterized by marked neuronal loss—seizure freedom was more often associated with preservation rather than resection of HFA-rich regions. Together, these findings suggest that while interictal HFA provides a generalizable biomarker for SOZ localization, its prognostic relevance is etiology-dependent, and surgical strategies prioritizing resection of HFA-involved areas may be ineffective or even counterproductive in encephalomalacia.

### Contribution of individual HFA features

HFA occurrence rate was among the most influential features for accurate SOZ classification (Figure 4), and clinician-defined SOZ sites were characterized by higher HFA occurrence rates than non-epileptic sites, with large effect sizes across all detectors (>0.7; **Table 1**; Figure 2). Elevated HFA occurrence rates localized SOZ sites across MRI-defined etiology groups in the external validation cohort, achieving AUROCs of 0.64–0.81 depending on the detector used (Figure 7A). This observation is consistent with the prevailing practice of using elevated occurrence rates to aid localization of the epileptogenic zone^12–14,17,45^.

Some investigators may question whether HFA occurrence rates alone are sufficient and whether additional features meaningfully contribute to localization of the epileptogenic zone. Our findings indicate that the extent to which morphological and anatomical features contribute to SOZ classification varies across HFA detectors. MNI-defined HFA appeared to benefit least from consideration of features beyond occurrence rate, whereas HFA defined by other detectors showed more substantial improvement when additional features were incorporated. Specifically, in the analysis of MNI-defined HFA, the model-derived SOZ probability score showed improved classification of clinician-defined SOZ compared with occurrence rate in only one geographical external validation cohort, but not in the remaining three cohorts (Figure 3D). The MNI detector is uniquely designed to be insensitive to prolonged high-frequency augmentations commonly observed in non-epileptic sensory cortices and hippocampi^48^. In contrast, for HIL-defined HFA, the probability score improved SOZ classification relative to occurrence rate in three of the four cohorts (Figure 3).

The next strongest contributors to SOZ classification were spectral entropy and spectral power, although their effect sizes varied across detectors. As illustrated by the SHAP dependence plot (Figure 4C), mid-range entropy values (∼0.7–0.8) were associated with a higher likelihood of SOZ sites, whereas both lower and higher entropy values contributed less to SOZ localization. HFA events with entropy values of ∼0.7–0.8 corresponded to broadband HFA associated with interictal spike discharges, whereas events with lower entropy values (∼0.5–0.6) reflected distinct HFOs characterized by a narrow spectral peak and minimal broadband activity (**Figure S1**). The observation that HFA events with mid-range entropy values were more likely to be associated with SOZ sites is consistent with a recent international multicenter study of 109 patients, which showed that the extent of resection of sites generating spike-associated HFA_80-250 Hz_ (so-called spike ripples)—rather than spikes or HFA events alone—provided more accurate prognostic information^17^. The XGBoost model demonstrated that the relationship between spectral entropy and seizure-onset likelihood was non-linear (Figure 4C), suggesting that such associations may not be readily captured by conventional linear statistical approaches.

SOZ sites exhibited higher spectral power than non-epileptic sites, with medium to large effect sizes (0.44–0.68) for STE-, SLL-, and HIL-defined HFA (**Table 1**; Figure 2). In contrast, MNI-defined HFA power did not significantly differ between SOZ and non-epileptic sites. The association between elevated HFA spectral power and increased seizure-onset likelihood is consistent with a prior study of 90 patients with drug-resistant focal epilepsy, which reported that spontaneously generated HFA at SOZ sites showed higher spectral power than task-induced HFA during cognitive and motor paradigms^20^. Conversely, a study of 10 patients reported comparable spectral power of spontaneously generated HFA between extra-occipital SOZ sites and non-epileptic primary visual cortex^49^.

Although HFA frequency band contributed to SOZ classification with variable effect sizes, depending on the detector (**Table 1**), the frequency ranges of HFA events largely overlapped between SOZ and non-epileptic sites (Figure 2D). Therefore, our findings support the notion that identifying HFA events with higher spectral frequency bands may not enhance localization accuracy of the epileptogenic zone^50^. A study of 109 patients reported that spike-associated HFA_80–250 Hz_ was more readily detected and more informative for epileptogenic zone localization than HFA_250–500 Hz_; specifically, approximately 80% of patients with ILAE Class 1 outcomes underwent inclusive resection of sites exhibiting spike-associated HFA_80–250 Hz_, whereas only 36% had inclusive resection of sites with HFA_250–500 Hz_^17^

Likewise, the durations of HFA events largely overlapped between SOZ and non-epileptic sites, except for SLL-defined HFA (Figure 2E). A prior study reported that the duration of spontaneously generated HFA at SOZ sites was 27 ± 23 ms (mean ± standard deviation), compared with 22 ± 19 ms for task-induced HFA^20^. Another study found that spontaneously generated HFA in non-epileptic primary visual cortex had longer durations than those generated by extra-occipital SOZ sites^49^.

### Etiology-dependent limitations of HFA in predicting postoperative seizure outcomes

In our derivation cohort, we demonstrated that a higher ’SOZ probability biomarker difference’—reflecting the preferential resection of HFA-rich regions while preserving HFA-poor regions—was associated with ILAE Class 1 (Figure 5A). Moreover, this biomarker improved the predictive performance of a prognostic model based on standard clinical variables (Figure 6B). However, this utility did not generalize to the three external validation cohorts (**Figures 5B–5D; 6C–6E**), suggesting limitations in the utility of SOZ probability score as a biomarker. The ’SOZ probability biomarker difference’ classified ILAE Class 1 outcomes with AUROC values ranging from 0.55–0.73 in non-lesional epilepsy but as low as 0.21–0.46 in encephalomalacia (Figure 7B). What explains the unexpected finding that patients with encephalomalacia had better surgical outcomes when HFA-rich regions were left intact rather than removed? One plausible explanation is that the epileptogenic zone in encephalomalacia often involves scarred tissue where cortical parenchyma is largely absent, leading to reduced generation of HFA signals compared to patients with preserved cortical architecture. Prior studies have suggested that, in encephalomalacia, chronic epileptogenicity may be primarily driven by impaired inhibitory mechanisms rather than excessive excitation^51,52^. It is possible that physiological HFA from preserved eloquent cortex was more prominent than pathological HFA from the scarred regions that were resected, resulting in a lower biomarker difference in ILAE Class 1 cases. Alternatively, technical limitations in intracranial electrode sampling—particularly at the interface between scarred and intact cortex—may have hindered accurate localization of epileptogenic HFA. These findings underscore the critical insight that the notion of “the more HFA-rich cortex resected, the better the seizure outcome” does not uniformly apply across MRI-defined etiologies.

### Methodological limitations

This retrospective study analyzed the first 5–20 minutes of non-REM sleep, yielding a lower level of evidence than prospective, registered investigations^14,18^. Within the three external validation cohorts (n = 91), higher SOZ probability biomarker difference did not predict ILAE Class 1 outcomes, with AUROCs of 0.46–0.54 (Figure 7B), indicating chance-level performance. The limited predictive performance of the SOZ probability score does not necessarily indicate a fundamental shortcoming of the ‘biomarker difference’ itself^34^. Even when using four other summary metrics to quantify the extent of HFA-rich tissue resection^17,45–47^, predictive accuracy remained poor in these external validation cohorts (**Figures S8-S11**). Although longer than some intraoperative HFA recordings^18^, our analysis epochs were substantially shorter than the multi-day recordings used by Dimakopoulos et al. (2025)^18^, who defined clinically meaningful HFA-rich areas only when HFA was reproducible across sessions and excluded patients lacking such reproducibility from the subsequent ROC analysis. In their cohort, complete HFA resection yielded an estimated AUROC of ∼0.61 for seizure freedom^18^. These findings underscore the need for prospective, objective definitions of clinically meaningful HFA-rich areas and for surgical strategies tailored to specific epilepsy etiology.

Our study also did not incorporate network-level analysis of HFA across distant electrode sites. Prior work suggests that the onset zone of HFA activity may better localize the epileptogenic zone^19,46^. Although regions with the highest HFA rates often coincide with the onset zone, this is not universally the case. Future studies should investigate whether incorporating inter-electrode HFA network features improves the external validity of HFA as a biomarker.

Finally, this study was conducted primarily in a patient population undergoing subdural electrode implantation, occasionally supplemented with depth electrodes. Neither stereoelectroencephalography nor high-density subdural grids^53^ were used. It remains unclear whether these alternative recording modalities might more effectively capture the epileptogenic zone, particularly in encephalomalacia. *Methodological* external validation using stereoelectroencephalography or high-density grids is warranted in future research.

## Data availability

Upon acceptance of the manuscript, the complete dataset will be made publicly available through OpenNeuro.

## Code availability

The codes are openly available on GitHub: https://github.com/KeHatano/comprehensive_HFA_based_model/tree/v1.0.0

## Funding

This work was supported by the National Institutes of Health (NIH; NS064033 to E.A.), the Uehara Memorial Foundation Postdoctoral Fellowship (202441017 to K.H.; 20210301 to H.U.), and the Japan Society for the Promotion of Science (JP22J23281 and JP22KJ0323 to N.K.; 202560628 to H.U.; JP19K09494 and 22K09296 to M.I.).

## Competing interests

The authors report no competing interests.

## Abbrevitations

ASM: antiseizure medication
AUC: areas under the curve
AUPRC: area under the precision-recall curve
AUROC: area under the receiver operating characteristic curve
HFA: high-frequency activity
HFO: high-frequency oscillation
HIL: Hilbert
iEEG: intracranial electroencephalography
ILAE: International League Against Epilepsy
IQR: interquartile range
MNI: Montreal Neurological Institute
NCNP: National Center of Neurology and Psychiatry
OR: odds ratio
SHAP: SHapley Additive exPlanations
SD: standard deviation
SLL: Short Line Length
SOZ: seizure onset zone
STE: Short Time Energy
XGBoost: eXtreme Gradient Boosting

## Notes

### Competing Interest Statement

The authors have declared no competing interest.

### Author Declarations

Ethics committee/IRB of Wayne State University (#048404MP2E), Tohoku University (#2023-1-1114-1), the National Center of Neurology and Psychiatry (#A2021-050) gave ethical approval for this work.

