## Supplementary Material for "Internal and External Validation of Comprehensive High-Frequency Activity Biomarkers for Epilepsy Surgery"

This supplementary document includes the following materials:

**Table S1. Hyperparameter search ranges and optimal values for the XGBoost classifier.**

**Table S2. Patient profile.**

**Table S3. Model performance for classifying pseudo-seizure onset zone (SOZ) sites.**

**Table S4. Seizure onset zone (SOZ) probability score and seizure outcome (univariate analysis).**

**Table S5. Seizure onset zone (SOZ) probability score and seizure outcome (multivariate analysis).**

**Table S6. High-frequency activity (HFA) rate and seizure outcome (univariate analysis).**

**Table S7. High-frequency activity (HFA) rate and seizure outcome (multivariate analysis).**

**Figure S1. Examples of spectral entropy in two types of high-frequency activity (HFA) events.**

**Figure S2. Summary measures reflecting the resection completeness.**

**Figure S3.** **Spatial distribution of electrode sites.**

**Figure S4. High-frequency activity (HFA) features across ages.**

**Figure S5. Precision of intracranial EEG biomarker–based classification of the seizure onset zone (SOZ).**

**Figure S6. Precision value of intracranial EEG biomarkers for classification of seizure outcomes.**

**Figure S7. Additive precision value of intracranial EEG biomarkers for classification of seizure outcomes.**

**Figure S8. Outcome prediction using ‘difference index’ (Shi et al., 2024).**

**Figure S9. Outcome prediction using ‘resection ratio’ (Nevalainen et al., 2020).**

**Figure S10. Outcome prediction using ‘critical resection percentage’ (Lin et al., 2024).**

**Figure S11. Outcome prediction using ‘distinguishability statistic’ (Taylor et al., 2022).**

| Hyperparameter | Search space | Optimal value | | | |
| --- | --- | --- | --- | --- | --- |
|  |  | STE | SLL | HIL | MNI |
| *learning_rate* | 0.01–0.3 | 0.0323 | 0.0498 | 0.047 | 0.07 |
| *max_depth* | 3–10 | 9 | 8 | 8 | 9 |
| *min_child_weight* | 1–10 | 1 | 1 | 1 | 1 |
| *subsample* | 0.5–1.0 | 0.9348 | 0.9343 | 0.9161 | 0.8776 |
| *colsample_bytree* | 0.5–1.0 | 0.761 | 0.8313 | 0.7821 | 0.8661 |
| *gamma* | 0–5 | 0.2031 | 0.0962 | 0.1899 | 0.1645 |
| *reg_alpha* | 0–5 | 0.5404 | 0.2577 | 0.6973 | 0.3482 |
| *reg_lambda* | 0–5 | 3.0278 | 2.4099 | 1.9089 | 1.4855 |

**Table S1. Hyperparameter search ranges and optimal values for the XGBoost classifier.** In the outer loop, the training datasets for SOZ classification model was divided into 10 folds using stratified group cross-validation, where age was binned into five groups (0–5, 5–10, 10–15, 15–20, and ≥20 years) to reduce age imbalance. In the inner loop, 10-fold cross-validation was performed at the electrode level on the training portion of each outer fold, to select the optimal hyperparameters based on the mean logarithmic loss (log loss), which quantified the accuracy of predicted probabilities. A hyperparameter search space was predefined for XGBoost, including *learning_rate*, *max_depth*, *min_child_weight*, *subsample*, *colsample_bytree*, *gamma*, *reg_alpha*, and *reg_lambda*. A total of 200 trials were conducted for each outer fold to explore the hyperparameter space. The final model was trained on the entire training datasets for SOZ classification model using the averaged optimal hyperparameters obtained from the 10 outer folds.

The optimal hyperparameter values for each high-frequency activity (HFA) detector are presented. STE: Short Time Energy method. SLL: Short Line Length method. HIL: Hilbert method. MNI: Montreal Neurological Institute method.

|  | Derivation cohort (Detroit Medical Center) | |  | Validation cohorts | | |
| --- | --- | --- | --- | --- | --- | --- |
|  | Training and internal  cross-validation  for the SOZ classification  (*n* = 79) | Training and  internal  cross-validation for the outcome classification  (*n* = 142) |  | Detroit  Medical  Center  (*n* = 35) | Tohoku University  (n = 26) | NCNP  (n = 30) |
| Age,  years old | 11.3 ± 7.4 (range: 0–41) | 10.5 ± 7.3  (range: 0–45) |  | 10.0 ± 5.1 (range: 0–20) | 25.7 ± 8.8 (range: 12–42) | 20.9 ± 11.5 (range: 5–45) |
| Sex (female), n | 38 (48.1 %) | 70 (49.3 %) |  | 17 (48.6 %) | 14 (53.8 %) | 13 (43.3 %) |
| Daily seizures, n | 34 (43.0 %) | 57 (40.1 %) |  | 10 (28.6 %) | 7 (26.9 %) | 7 (23.3 %) |
| Number of ASMs | 2.1 ± 0.8 (range: 1–5) | 2.1 ± 0.8  (range: 1–5) |  | 2.3 ± 0.9 (range: 1–5) | 3.1 ± 0.9 (range: 1–5) | 2.7 ± 1.0 (range: 1–5) |
| Left-hemispheric epilepsy, n | 42 (53.2 %) | 73 (51.4 %) |  | 13 (37.1 %) | 15 (57.7 %) | 14 (46.7 %) |
| MRI-visible lesion, n | 52 (65.8 %) | 89 (62.7 %) |  | 25 (71.4 %) | 16 (61.5 %) | 24 (80 %) |
| Habitual seizures captured during iEEG recording, n | 79 (100 %) | 127 (89.4 %) |  | 31 (88.6 %) | 26 (100 %) | 30 (100 %) |
| Incomplete resection  of SOZ, n | 6 (7.6 %) | 29 (20.4 %) |  | 3 (8.6 %) | 6 (23.1 %) | 18 (60 %) |
| Necessity to resect extra-temporal region, n | 49 (62.0 %) | 95 (66.9 %) |  | 30 (85.7 %) | 11 (42.3 %) | 20 (66.7 %) |
| Resection extent (%) | 18.9 ± 23.6 (range:  0.6–89.8) | 21.3 ± 24.7  (range: 0.1–91.6) |  | 23.7 ± 21.7 (range:  1.6–84.7) | 4.9 ± 3.4 (range:  0.8–13.0) | 4.7 ± 3.7 (range:  0.3–15.4) |
| ILAE surgical outcome, n |  |  |  |  |  |  |
| Class 1 | 79 (100 %) | 90 (63.4 %) |  | 19 (54.3 %) | 16 (61.5 %) | 13 (43.3 %) |
| Class 2 | 0 (0 %) | 4 (2.8 %) |  | 0 (0 %) | 1 (3.8 %) | 3 (10 %) |
| Class 3 | 0 (0 %) | 15 (10.6 %) |  | 3 (8.6 %) | 4 (15.4 %) | 1 (3.3 %) |
| Class 4 | 0 (0 %) | 24 (16.9 %) |  | 9 (25.7 %) | 1 (3.8 %) | 7 (23.3 %) |
| Class 5 | 0 (0 %) | 9 (6.3 %) |  | 4 (11.4 %) | 4 (15.4 %) | 5 (16.7 %) |
| Class 6 | 0 (0 %) | 0 (0 %) |  | 0 (0 %) | 0 (0 %) | 1 (3.3 %) |
| Electrode number |  |  |  |  |  |  |
| Total | 4,905 * | 15,464 |  | 3,934 | 1,644 | 1,897 |
| Frontal lobe other than central area | 1,258 (left: 627, right: 631) | 4,208 (left: 1992,  right: 2216) |  | 1,100 (left: 294, right: 806) | 353 (left: 206, right: 147) | 394 (left: 152, right: 242) |
| Central area | 1,000 (left: 502, right: 498) | 2,935 (left: 1476,  right: 1459) |  | 676 (left: 240, right: 436) | 226 (left: 127, right: 99) | 219 (left: 104, right: 115) |
| Parietal lobe other than central area | 715 (left: 352, right: 363) | 2,387 (left: 1069,  right: 1318) |  | 693 (left: 255, right: 438) | 284 (left: 129, right: 155) | 385 (left: 176, right: 209) |
| Occipital lobe | 398 (left: 225, right: 173) | 1,390 (left: 695,  right: 695) |  | 349 (left: 126, right: 223) | 95 (left: 48, right: 47) | 161 (left: 67, right: 94) |
| Temporal lobe | 1,534 (left: 958, right: 576) | 4,543 (left: 2421,  right: 2122) |  | 1,103 (left: 424, right: 679) | 686 (left: 437, right: 249) | 738 (left: 456, right: 282) |
| Insula | 0 | 1 (left: 1, right: 0) |  | 13 (left: 6, right: 7) | 0 (left: 0, right: 0) | 0 (left: 0, right: 0) |
| Depth electrode sites | 6 | 13 |  | 224 | 18 | 66 |
| Preoperative MRI findings |  |  |  |  |  |  |
| Non-lesion, n | 27 (34.2 %) | 53 (37.3 %) |  | 10 (28.6 %) | 10 (38.5 %) | 6 (20 %) |
| Dysplasia, n | 25 (31.6 %) | 45 (31.7 %) |  | 11 (31.4 %) | 7 (26.9 %) | 10 (33.3 %) |
| Hippocampal sclerosis, n | 2 (2.5 %) | 3 (2.1 %) |  | 0 (0 %) | 3 (11.5 %) | 0 (0 %) |
| Tumor, n | 13 (16.5 %) | 21 (14.8 %) |  | 8 (22.9 %) | 4 (15.4 %) | 1 (3.3 %) |
| Encephalomalacia, n | 6 (7.6 %) | 7 (4.9 %) |  | 3 (8.6 %) | 0 (0 %) | 7 (23.3 %) |
| Others, n | 7 (8.9 %) | 15 (10.6 %) |  | 4 (11.4 %) | 4 (15.4 %) | 6 (20 %) |

**Table S2. Patient profile.** ASM: anti-seizure medication taken immediately before the electrode placement. iEEG: intracranial EEG; ILAE: International League Against Epilepsy. MRI: magnetic resonance imaging. SOZ: seizure-onset zone. *: Of the 4,905 electrode sites, 822 were SOZ and 4,083 were non-epileptic.

|  | Mean ± SD | 95%CI |
| --- | --- | --- |
| STE | 0.50 ± 0.018 | 0.47–0.53 |
| SLL | 0.50 ± 0.017 | 0.47–0.53 |
| HIL | 0.50 ± 0.016 | 0.47–0.55 |
| MNI | 0.50 ± 0.020 | 0.46–0.54 |

**Table S3. Model performance for classifying pseudo-seizure onset zone (SOZ) sites.**

This table summarizes the nested cross-validation area under the receiver operating characteristic curve (AUROC) for classifying pseudo-SOZ sites in the derivation cohort for the SOZ classification. Across all HFA detectors, AUROC values did not differ from 0.5 based on the permutation test. STE detector: Short Time Energy detector. SLL: Short Line Length. HIL: Hilbert. MNI: Montreal Neurological Institute.

|  | STE | |  | SLL | |  | HIL | |  | MNI | |
| --- | --- | --- | --- | --- | --- | --- | --- | --- | --- | --- | --- |
|  | Odds ratio (95%CI) | p-value |  | Odds ratio (95%CI) | p-value |  | Odds ratio (95%CI) | p-value |  | Odds ratio (95%CI) | p-value |
| Biomarker difference of SOZ probability | 24.2 (3.73–157.06) | 0.001 |  | 36.88 (5.33–255.09) | <0.001 |  | 20.72 (3.46–123.96) | 0.001 |  | 9.77 (2.09–45.57) | 0.004 |

**Table S4. Seizure onset zone (SOZ) probability score and seizure outcome (univariate analysis).**

This table summarizes the effect of more complete resection of electrode sites with elevated SOZ probability scores on postoperative seizure outcome, as estimated using univariate logistic regression in the 142-patient derivation cohort. The SOZ probability score was derived from the comprehensive HFA-based model. Resection completeness was quantified using the ‘biomarker difference’ summary measure (**Figure S2A**). A larger biomarker difference reflects preferential resection of sites exhibiting higher SOZ probability scores, with relative preservation of sites showing lower SOZ probability scores. The effect size is presented as an odds ratio (with 95% CI), indicating the change in odds of achieving an ILAE class 1 outcome for each one-unit increase in the biomarker difference for a given patient. STE detector: Short Time Energy detector. SLL: Short Line Length. HIL: Hilbert. MNI: Montreal Neurological Institute.

|  | STE | |  | SLL | |  | HIL | |  | MNI | |
| --- | --- | --- | --- | --- | --- | --- | --- | --- | --- | --- | --- |
|  | Odds ratio (95%CI) | p-value |  | Odds ratio (95%CI) | p-value |  | Odds ratio (95%CI) | p-value |  | Odds ratio (95%CI) | p-value |
| Age (years old) | 1.0 (0.94–1.06) | 0.965 |  | 0.99 (0.93–1.05) | 0.761 |  | 1.0 (0.94–1.07) | 0.907 |  | 1.0 (0.94–1.06) | 0.949 |
| Sex (female) | 1.01 (0.46–2.23) | 0.986 |  | 1.15 (0.51–2.60) | 0.737 |  | 1.03 (0.47–2.29) | 0.936 |  | 1.07 (0.48–2.34) | 0.875 |
| Daily seizures | 2.8 (1.04–7.59) | 0.042 |  | 3.18 (1.14–8.83) | 0.027 |  | 2.89 (1.06–7.91) | 0.039 |  | 2.71 (1.02–7.21) | 0.046 |
| Number of anti-seizure medications | 0.67 (0.40–1.12) | 0.129 |  | 0.6 (0.35–1.02) | 0.061 |  | 0.66 (0.39–1.10) | 0.108 |  | 0.67 (0.40–1.11) | 0.119 |
| Left-hemispheric epilepsy | 0.57 (0.25–1.33) | 0.193 |  | 0.59 (0.25–1.39) | 0.227 |  | 0.55 (0.24–1.28) | 0.167 |  | 0.6 (0.27–1.37) | 0.228 |
| MRI-visible lesion | 0.83 (0.35–1.97) | 0.673 |  | 0.86 (0.36–2.07) | 0.734 |  | 0.87 (0.36–2.08) | 0.755 |  | 0.85 (0.36–2.00) | 0.708 |
| Habitual seizures captured during iEEG recording | 0.42 (0.11–1.67) | 0.22 |  | 0.38 (0.09–1.55) | 0.179 |  | 0.4 (0.10–1.58) | 0.192 |  | 0.45 (0.11–1.77) | 0.251 |
| Incomplete resection of seizure onset zone | 0.15 (0.05–0.47) | 0.001 |  | 0.14 (0.04–0.46) | 0.001 |  | 0.14 (0.05–0.44) | 0.001 |  | 0.16 (0.05–0.47) | 0.001 |
| Necessity to resect extra-temporal region | 1.24 (0.46–3.37) | 0.669 |  | 1.28 (0.47–3.52) | 0.628 |  | 1.15 (0.43–3.10) | 0.775 |  | 1.15 (0.43–3.08) | 0.778 |
| Resection extent (%) | 0.99 (0.97–1.01) | 0.183 |  | 0.98 (0.97–1.00) | 0.132 |  | 0.99 (0.97–1.01) | 0.249 |  | 0.99 (0.97–1.01) | 0.216 |
| Biomarker difference of SOZ probability | 34.33 (3.71–317.83) | 0.002 |  | 87.45 (8.10–944.49) | <0.001 |  | 31.7 (3.90–257.38) | 0.001 |  | 11.3 (1.81–70.62) | 0.01 |

**Table S5. Seizure onset zone (SOZ) probability score and seizure outcome (multivariate analysis).**

This table summarizes the additive effect of more complete resection of electrode sites with elevated SOZ probability scores on postoperative seizure outcome, as assessed by multivariate logistic regression incorporating 10 routinely available clinical predictors, in the 142-patient derivation cohort. The SOZ probability score was derived from the comprehensive HFA-based model. Resection completeness was quantified using the ‘biomarker difference’ summary measure (**Figure S2A**). A larger biomarker difference reflects preferential resection of sites exhibiting higher SOZ probability scores, with relative preservation of sites showing lower SOZ probability scores. The effect size is presented as an odds ratio (with 95% CI), indicating the change in odds of achieving an ILAE class 1 outcome for each one-unit increase in the biomarker difference for a given patient. STE detector: Short Time Energy detector. SLL: Short Line Length. HIL: Hilbert. MNI: Montreal Neurological Institute.

|  | STE | |  | SLL | |  | HIL | |  | MNI | |
| --- | --- | --- | --- | --- | --- | --- | --- | --- | --- | --- | --- |
|  | Odds ratio (95%CI) | p-value |  | Odds ratio (95%CI) | p-value |  | Odds ratio (95%CI) | p-value |  | Odds ratio (95%CI) | p-value |
| Biomarker difference of HFA rate | 1.72 (0.85–3.48) | 0.132 |  | 1.04 (0.94–1.14) | 0.438 |  | 1.14 (0.95–1.37) | 0.167 |  | 1.69 (0.97–2.93) | 0.063 |

**Table S6. High-frequency activity (HFA) rate and seizure outcome (univariate analysis).**

This table summarizes the effect of more complete resection of electrode sites with elevated HFA rate on postoperative seizure outcome, as estimated using univariate logistic regression in the 142-patient derivation cohort. Resection completeness was quantified using the ‘biomarker difference’ summary measure (**Figure S2A**). A larger biomarker difference reflects preferential resection of sites exhibiting higher HFA rates, with relative preservation of sites showing lower HFA rates. The effect size is presented as an odds ratio (with 95% CI), indicating the change in odds of achieving an ILAE class 1 outcome for each one-unit increase in the biomarker difference for a given patient. STE detector: Short Time Energy detector. SLL: Short Line Length. HIL: Hilbert. MNI: Montreal Neurological Institute.

|  | STE | |  | SLL | |  | HIL | |  | MNI | |
| --- | --- | --- | --- | --- | --- | --- | --- | --- | --- | --- | --- |
|  | Odds ratio (95%CI) | p-value |  | Odds ratio (95%CI) | p-value |  | Odds ratio (95%CI) | p-value |  | Odds ratio (95%CI) | p-value |
| Age (years old) | 1.00 (0.94–1.06) | 0.908 |  | 0.99 (0.94–1.06) | 0.848 |  | 0.99 (0.93–1.06) | 0.825 |  | 1.00 (0.94–1.06) | 0.993 |
| Sex (female) | 1.03 (0.47–2.24) | 0.946 |  | 1.00 (0.46–2.17) | 0.999 |  | 0.93 (0.43–2.04) | 0.858 |  | 1.02 (0.46–2.22) | 0.966 |
| Daily seizures | 2.42 (0.91–6.48) | 0.078 |  | 2.18 (0.84–5.67) | 0.11 |  | 2.5 (0.93–6.74) | 0.07 |  | 2.50 (0.93–6.74) | 0.069 |
| Number of anti-seizure medications | 0.70 (0.43–1.16) | 0.166 |  | 0.71 (0.43–1.16) | 0.173 |  | 0.67 (0.41–1.11) | 0.122 |  | 0.72 (0.43–1.18) | 0.192 |
| Left-hemispheric epilepsy | 0.52 (0.22–1.20) | 0.126 |  | 0.51 (0.22–1.19) | 0.12 |  | 0.54 (0.23–1.25) | 0.15 |  | 0.52 (0.23–1.21) | 0.129 |
| MRI-visible lesion | 1.07 (0.46–2.51) | 0.875 |  | 1.09 (0.46–2.55) | 0.847 |  | 1.12 (0.48–2.65) | 0.79 |  | 1.03 (0.44–2.40) | 0.953 |
| Habitual seizures captured during iEEG recording | 0.85 (0.23–3.14) | 0.804 |  | 0.81 (0.22–3.07) | 0.761 |  | 0.79 (0.21–2.93) | 0.724 |  | 1.09 (0.29–4.08) | 0.902 |
| Incomplete resection of seizure onset zone | 0.10 (0.03–0.32) | <0.001 |  | 0.10 (0.03–0.33) | <0.001 |  | 0.09 (0.03–0.31) | <0.001 |  | 0.10 (0.03–0.33) | <0.001 |
| Necessity to resect extra-temporal region | 0.73 (0.28–1.88) | 0.511 |  | 0.85 (0.33–2.18) | 0.74 |  | 0.62 (0.24–1.64) | 0.335 |  | 0.74 (0.29–1.91) | 0.537 |
| Resection extent (%) | 0.99 (0.97–1.01) | 0.266 |  | 0.99 (0.97–1.01) | 0.215 |  | 0.99 (0.97–1.01) | 0.333 |  | 0.99 (0.97–1.01) | 0.33 |
| Biomarker difference of HFA rate | 2.86 (1.20–6.83) | 0.018 |  | 1.12 (0.99–1.27) | 0.062 |  | 1.36 (1.07–1.73) | 0.013 |  | 2.13 (1.17–3.85) | 0.013 |

**Table S7. High-frequency activity (HFA) rate and seizure outcome (multivariate analysis).**

This table summarizes the additive effect of more complete resection of electrode sites with elevated HFA rates on postoperative seizure outcome, as assessed by multivariate logistic regression incorporating 10 routinely available clinical predictors, in the 142-patient derivation cohort. Resection completeness was quantified using the ‘biomarker difference’ summary measure (**Figure S2A**). A larger biomarker difference reflects preferential resection of sites exhibiting higher HFA rates, with relative preservation of sites showing lower HFA rates. The effect size is presented as an odds ratio (with 95% CI), indicating the change in odds of achieving an ILAE class 1 outcome for each one-unit increase in the biomarker difference for a given patient. STE detector: Short Time Energy detector. SLL: Short Line Length. HIL: Hilbert. MNI: Montreal Neurological Institute.

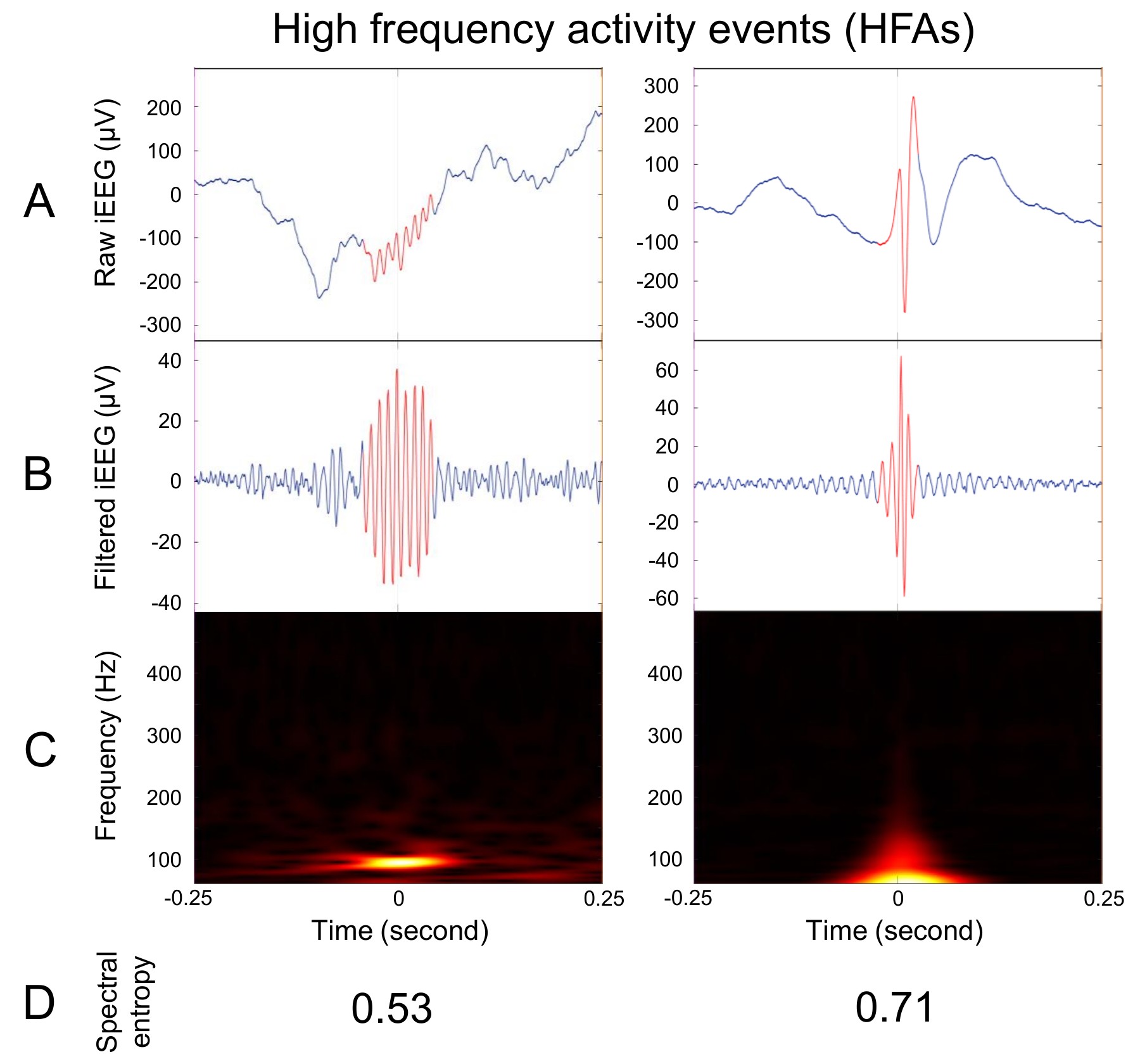

**Figure S1. Examples of spectral entropy in two types of high-frequency activity (HFA) events.** Left: An HFA event with power augmentation concentrated within a narrow frequency band (consistent with high-frequency oscillations, HFOs). Right: An HFA event with broadband power augmentation accompanying an interictal spike discharge. **A.** Raw intracranial EEG (iEEG). **B.** Band-pass filtered iEEG (80–500 Hz). Red segments indicate periods identified as HFA by RIPPLELAB. **C.** Time–frequency plots. **D.** Normalized spectral entropy values.

A. $\text{'Biomarker difference'}=\frac{\sum_{\text{Resected channels}} value}{N_{\text{Resected channels}}}-\frac{\sum_{\text{Preserved channels}} value}{N_{\text{Preserved channels}}}$
B. $\text{'Difference index'}=\frac{\sum_{\text{Resected channels}} value-\sum_{\text{Preserved channels}} value}{\sum_{\text{All }\text{channels}} value}$
C. $\text{'Resection ratio'}=\frac{\sum_{\text{Resected channels}} value*}{\sum_{\text{All}\text{ channels}} value*}$
D. $\text{'Critical resection percentage'}=\frac{N_{\text{Resected channels showing critically abnormal values}}}{N_{\text{All channels showing critically abnormal values}}}$
E. $\text{'Distinguishability statistic'}=\frac{U_{\text{Resected channels}}}{N_{\text{Resected channels}}\times N_{\text{Preserved channels}}}$

**Figure S2. Summary measures reflecting the resection completeness.** In the present study, we determined the utility of outcome classification models incorporating each of the following summary measures, referred to as ‘biomarker difference’ (Kuroda et al., 2025), ‘difference index’ (Shi et al., 2024), ‘resection ratio’ (Nevalainen et al., 2020), ‘critical resection percentage’ (Lin et al., 2024), and ‘distinguishability statistic’ (Taylor et al., 2022). A higher value generally indicates that cortical regions with markedly elevated biomarker values were more extensively included in the resection. **A.** ‘Biomarker difference’ was defined as the difference between the mean biomarker value across resected electrode sites and that across preserved sites. **B.** ‘Difference index’ was defined as the difference between the summed biomarker values across resected and preserved electrode sites, normalized by the total biomarker sum across all sites. **C.** ‘Resection ratio’ was defined as the sum of biomarker values across resected electrode sites, normalized by the total biomarker sum across all sites. *: Only values exceeding a predefined cutoff were included in the calculation of ‘resection ratio’, following the approach of Nevalainen et al. (2020). In the present study, the cutoff value was determined based on the highest Youden’s index derived from receiver operating characteristic (ROC) analysis used to classify seizure onset zone (SOZ) versus non-epileptic electrode sites in the derivation cohort. **D.** ‘Critical resection percentage’ was defined as the percentage of regions showing critically abnormal values that were included in the resection. In the present study, electrode sites ranked within the top 30% were defined as showing critically abnormal values, following the approach described by Lin et al. (2024). **E.** ‘Distinguishability statistic’ was defined as the Mann–Whitney U statistic comparing biomarker values between resected and preserved electrode sites, normalized by the number of possible resected–preserved electrode pairs.

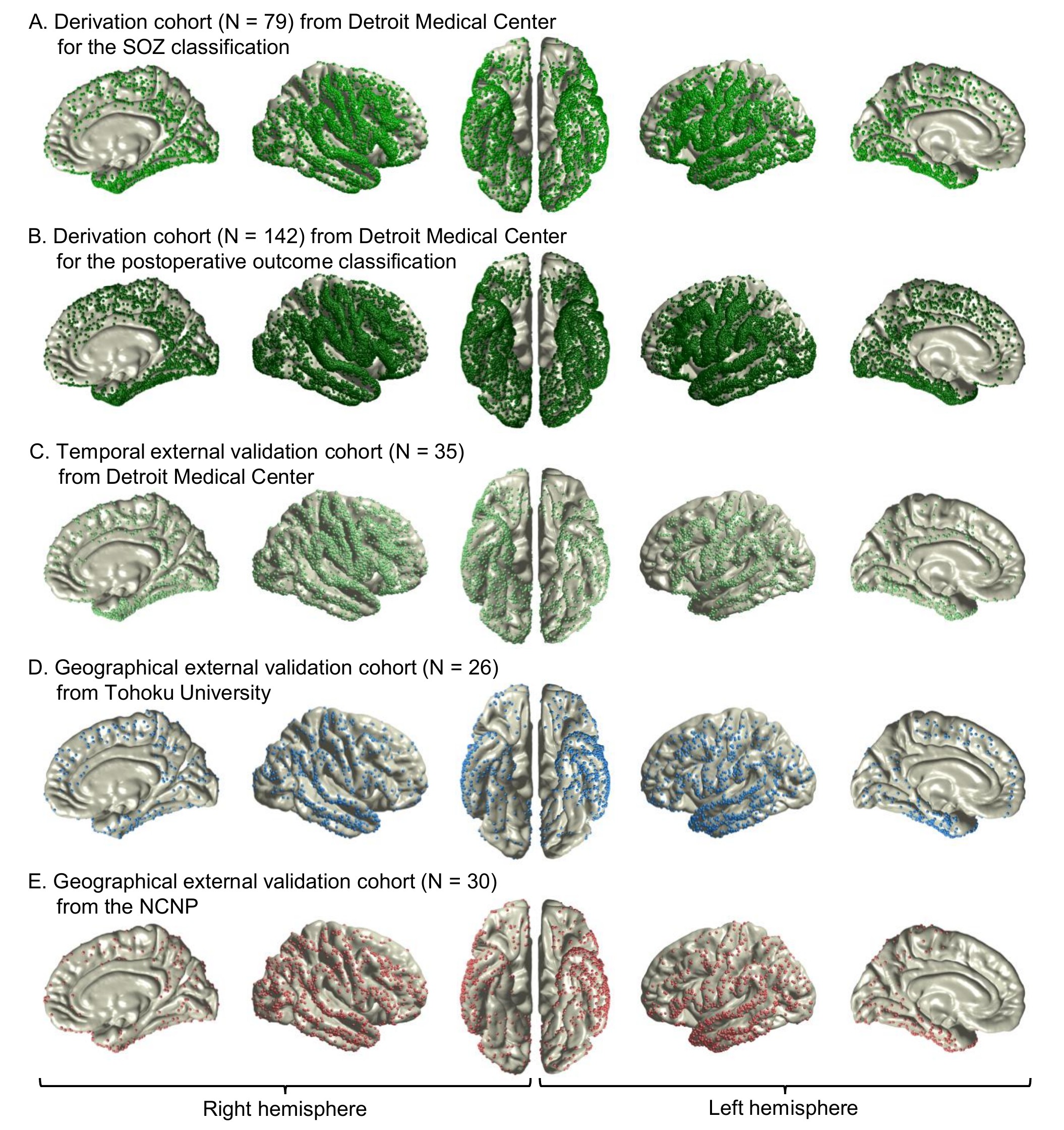

**Figure S3. Spatial distribution of electrode sites.**

**A.** Derivation cohort (N = 79) from Detroit Medical Center, used for training and internal cross-validation of the SOZ versus non-epileptic site classification model.
**B.** Derivation cohort (N = 142) from Detroit Medical Center, used for training and internal cross-validation of the postoperative outcome classification model.
**C.** Temporal external validation cohort (N = 35) from Detroit Medical Center, recruited subsequently to the derivation cohort, and used to evaluate SOZ- and outcome-classification performance.
**D.** Geographical external validation cohort (N = 26) from Tohoku University.
**E.** Geographical external validation cohort (N = 30) from the National Center of Neurology and Psychiatry (NCNP).

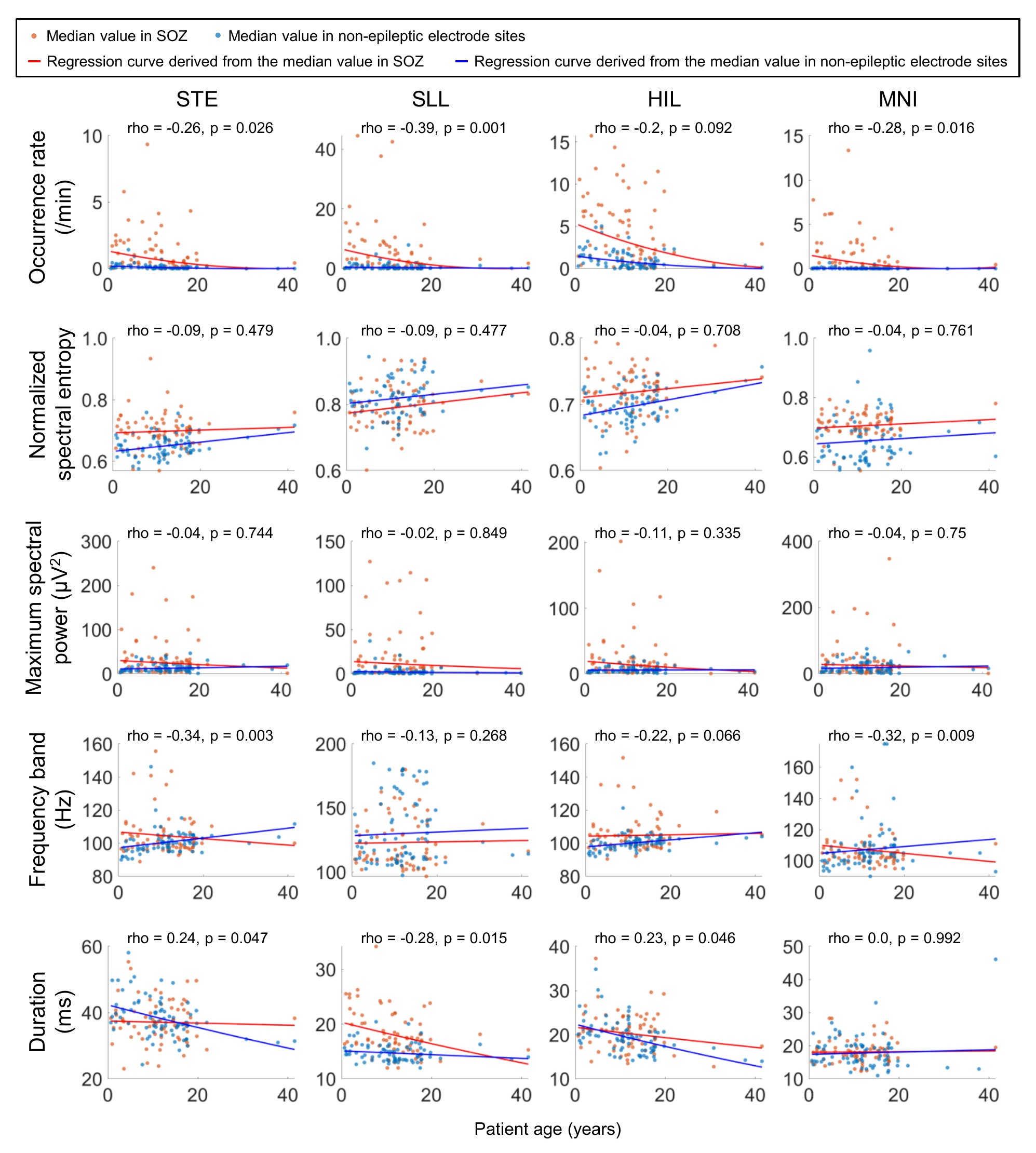

**Figure S4. High-frequency activity (HFA) features across ages.**

Scatter plots show HFA feature values (medians computed per patient) at seizure onset zone (SOZ; red) and non-epileptic sites (blue) across age among the 79 patients in the derivation cohort (see **Table S2**). Separate regression lines were fitted to each group using square-root–transformed HFA feature values. The difference between SOZ and non-epileptic values (SOZ – non-epileptic) was correlated with age, assessed using Spearman’s rho. **A.** Occurrence rate. **B.** Normalized spectral entropy. **C.** Maximum spectral power. **D.** Frequency band. **E.** Duration. A Spearman’s rho value greater than zero indicates that the SOZ–non-epileptic difference increased with patient age. STE detector: Short Time Energy detector. SLL: Short Line Length. HIL: Hilbert. MNI: Montreal Neurological Institute.

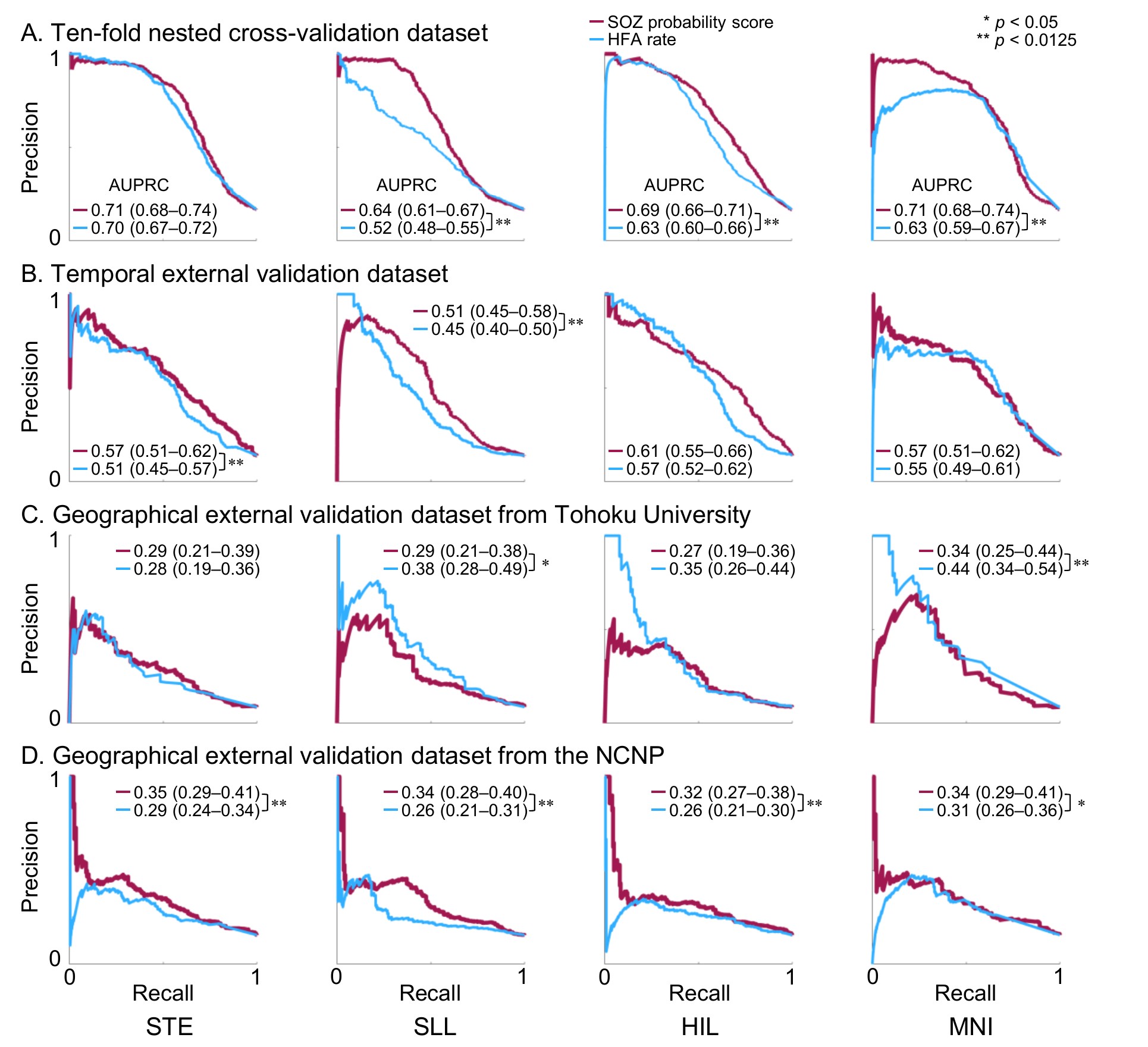

**Figure S5. Precision of intracranial EEG biomarker–based classification of the seizure onset zone (SOZ).**

Classification performance for distinguishing SOZ from non-epileptic sites using the model-derived SOZ probability score (red) and the high-frequency activity (HFA) occurrence rate alone (blue), quantified by the area under the precision-recall curve (AUPRC).

**A.** Ten-fold nested cross-validation within the internal cohort.

**B.** Temporal external validation dataset.

**C.** Geographical external validation dataset from Tohoku University.

**D.** Geographical external validation dataset from the National Center of Neurology and Psychiatry (NCNP). STE detector: Short-Time Energy detector. SLL: Short Line Length. HIL: Hilbert. MNI: Montreal Neurological Institute. * p < 0.05; ** p < 0.0125 derived from the DeLong test.

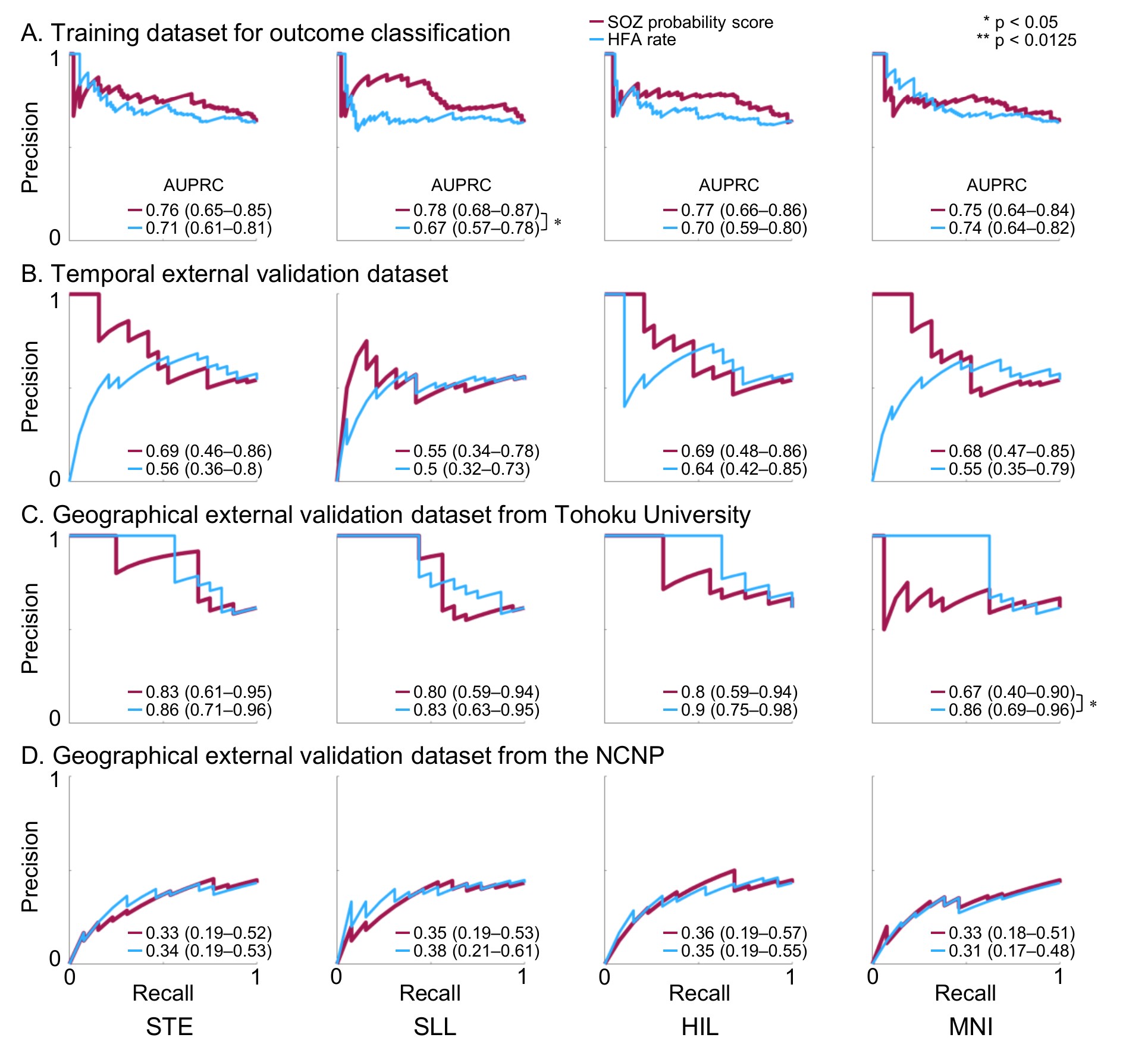

**Figure S6. Precision value of intracranial EEG biomarkers for classification of seizure outcomes.** Precision–recall curves illustrate the performance of classifying patients who achieved an ILAE class 1 outcome using the ‘seizure onset zone (SOZ) probability biomarker difference’ (red) and the ‘high-frequency activity (HFA) rate biomarker difference’ (blue). For each curve, the area under the precision–recall curve (AUPRC) with its 95% confidence interval (95% CI) is shown. STE detector: Short-Time Energy detector. SLL: Short Line Length. HIL: Hilbert. MNI: Montreal Neurological Institute. * p < 0.05; ** p < 0.0125 derived from the DeLong test.

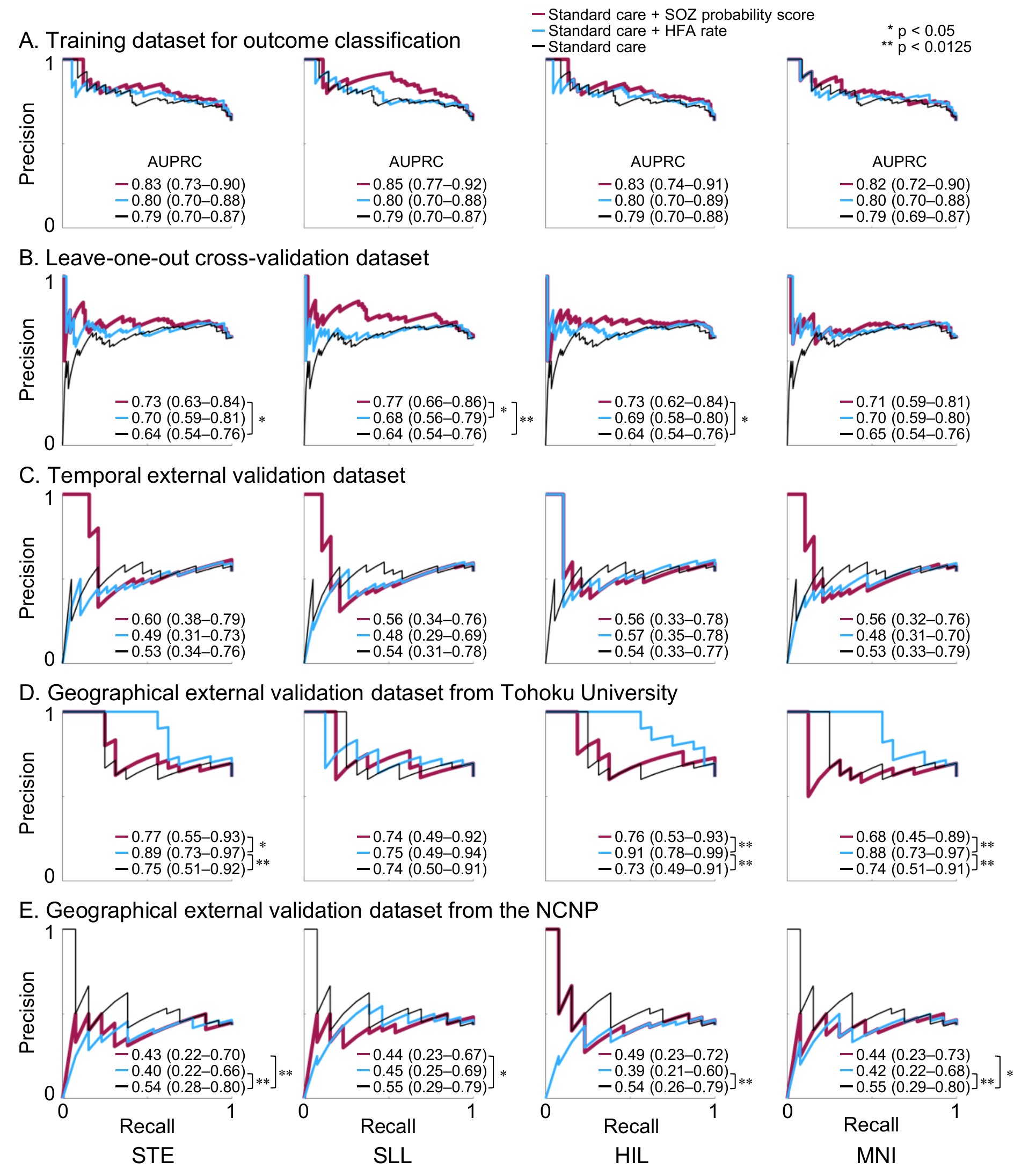

**Figure S7. Additive precision value of intracranial EEG biomarkers for classification of seizure outcomes.** Precision–recall curves illustrate the performance of three logistic regression models for classifying patients who achieved an ILAE class 1 outcome. The red curve represents the model incorporating the ‘seizure onset zone (SOZ) probability biomarker difference’ in addition to the standard-care model. The blue curve represents the model incorporating the ‘high-frequency activity (HFA) rate biomarker difference’ in addition to the standard-care model. The black curve represents the standard-care model alone. For each curve, the area under the precision–recall curve (AUPRC) with its 95% confidence interval (95% CI) is shown. STE detector: Short-Time Energy detector. SLL: Short Line Length. HIL: Hilbert. MNI: Montreal Neurological Institute. * p < 0.05; ** p < 0.0125 derived from the DeLong test.

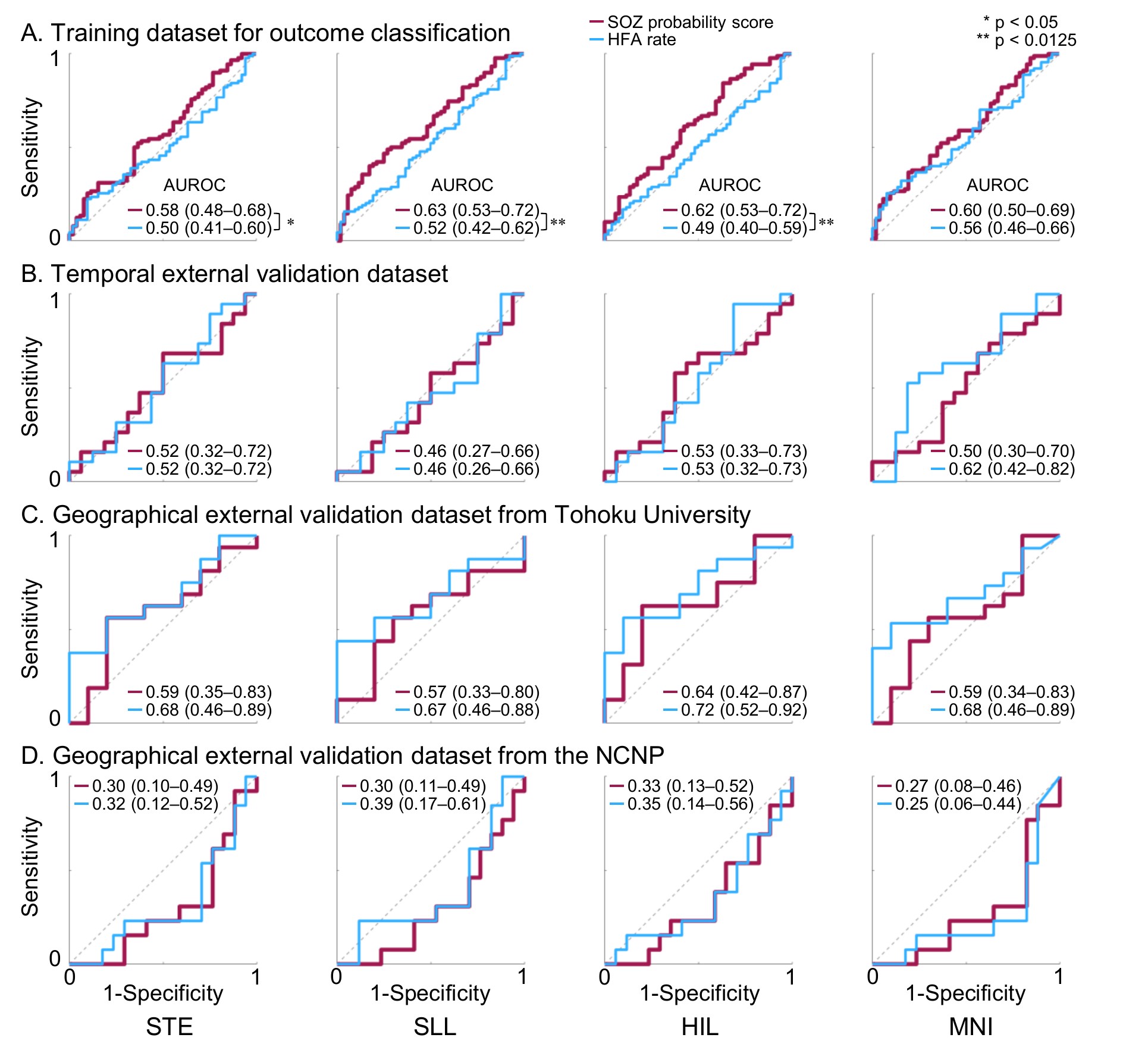

**Figure S8. Outcome prediction using ‘difference index’ (Shi et al., 2024).**

Receiver operating characteristic (ROC) curves present classification performance for classifying patients who achieved an ILAE class 1 outcome using the ‘seizure onset zone (SOZ) probability difference index’ (red) and the ‘high-frequency activity (HFA) rate difference index’ (blue). For each curve, the area under the ROC curve (AUROC) with its 95% confidence interval (95% CI) is shown.

**A.** Training dataset for outcome classification.

**B.** Temporal external validation dataset.

**C.** Geographical external validation dataset from Tohoku University.

**D.** Geographical external validation dataset from the National Center of Neurology and Psychiatry (NCNP). STE detector: Short-Time Energy detector. SLL: Short Line Length. HIL: Hilbert. MNI: Montreal Neurological Institute. * p < 0.05; ** p < 0.0125 derived from the DeLong test.

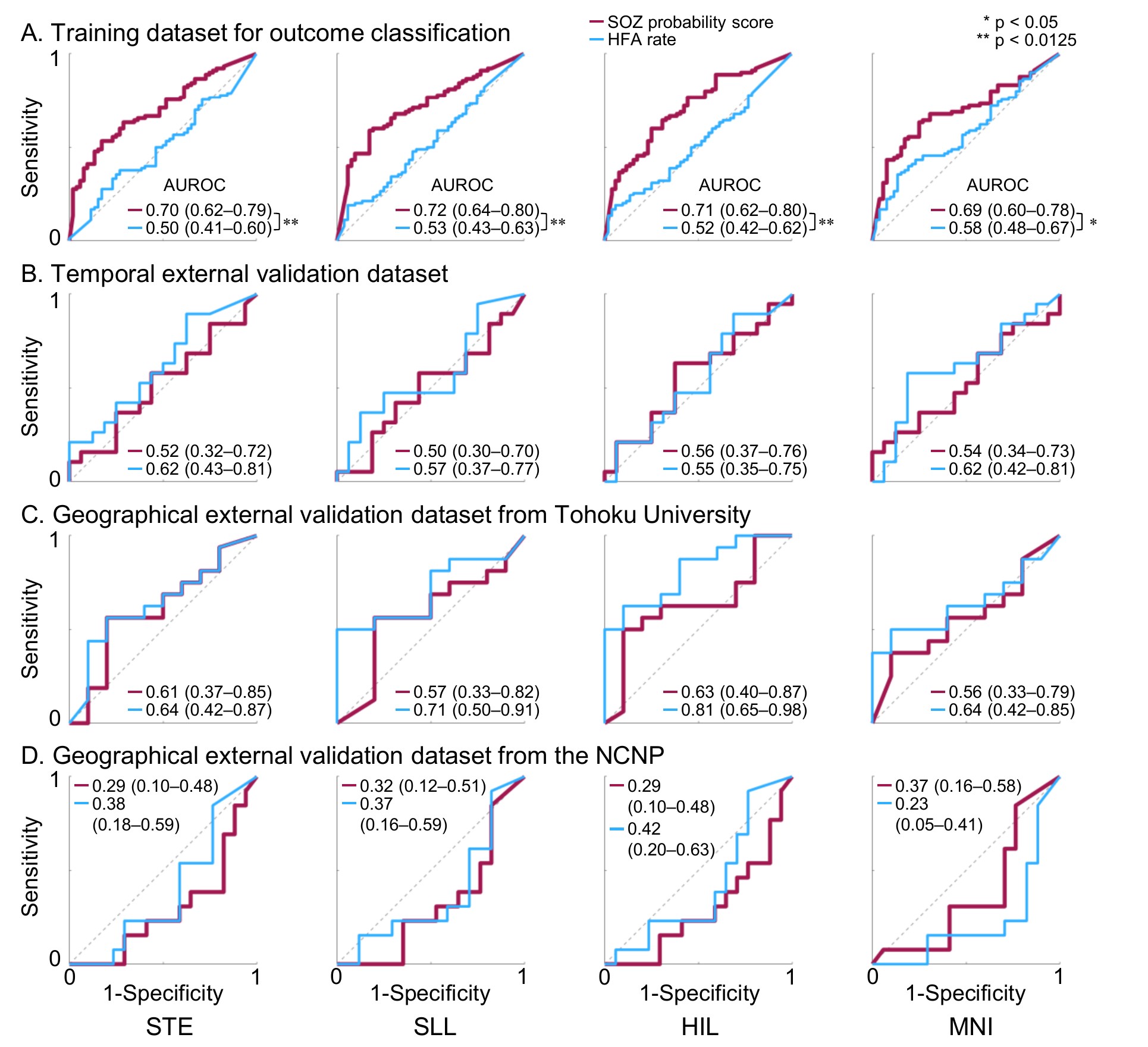

**Figure S9. Outcome prediction using ‘resection ratio’ (Nevalainen et al., 2020).**

Receiver operating characteristic (ROC) curves present classification performance for classifying patients who achieved an ILAE class 1 outcome using the ‘seizure onset zone (SOZ) probability resection ratio’ (red) and the ‘high-frequency activity (HFA) rate resection ratio’ (blue). For each curve, the area under the ROC curve (AUROC) with its 95% confidence interval (95% CI) is shown.

**A.** Training dataset for outcome classification.

**B.** Temporal external validation dataset.

**C.** Geographical external validation dataset from Tohoku University.

**D.** Geographical external validation dataset from the National Center of Neurology and Psychiatry (NCNP). STE detector: Short-Time Energy detector. SLL: Short Line Length. HIL: Hilbert. MNI: Montreal Neurological Institute. * p < 0.05; ** p < 0.0125 derived from the DeLong test.

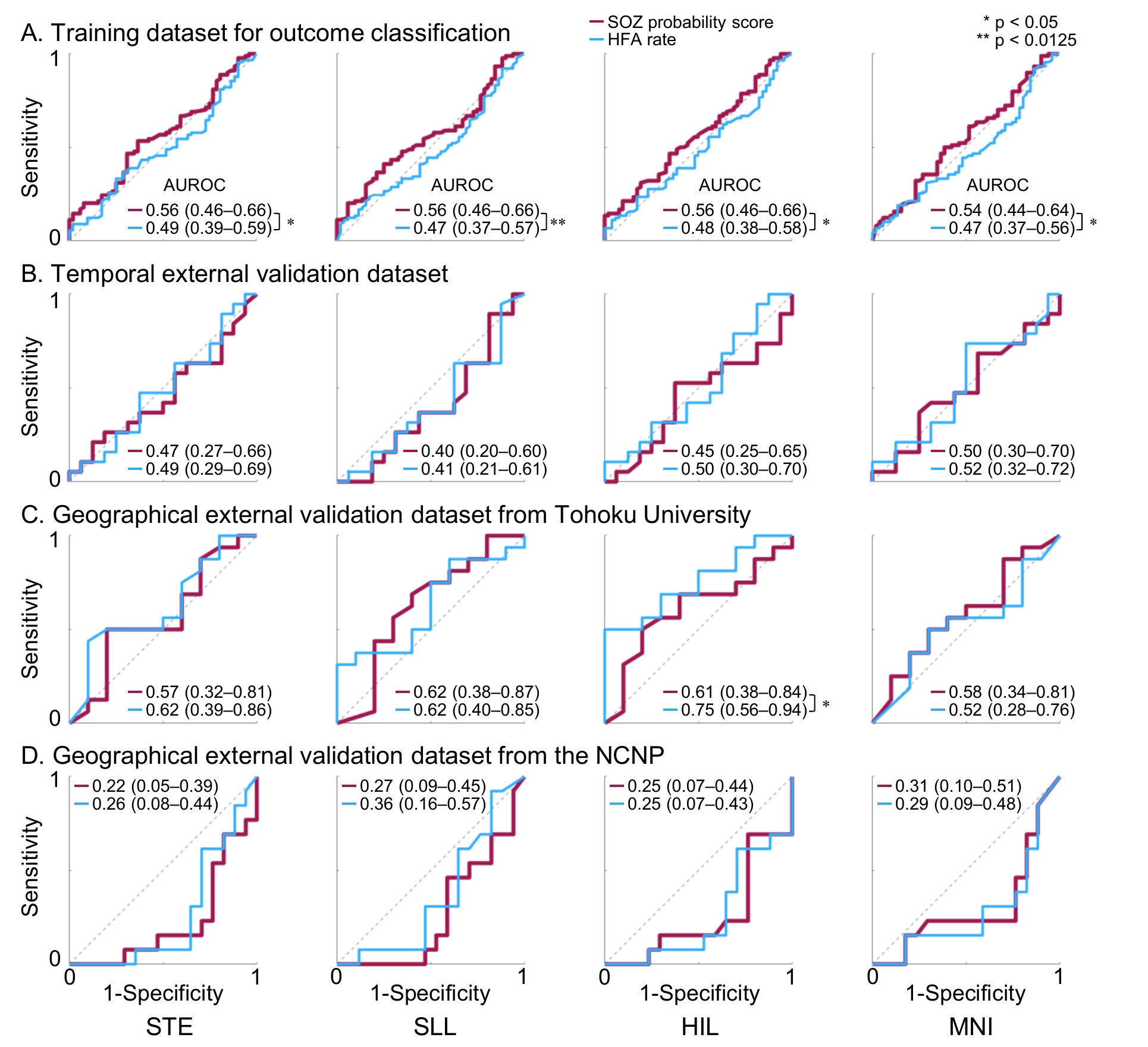

**Figure S10. Outcome prediction using ‘critical resection percentage’ (Lin et al., 2024).**

Receiver operating characteristic (ROC) curves present classification performance for classifying patients who achieved an ILAE class 1 outcome using the ‘seizure onset zone (SOZ) probability critical resection percentage’ (red) and the ‘high-frequency activity (HFA) rate critical resection percentage’ (blue). For each curve, the area under the ROC curve (AUROC) with its 95% confidence interval (95% CI) is shown.

**A.** Training dataset for outcome classification.

**B.** Temporal external validation dataset.

**C.** Geographical external validation dataset from Tohoku University.

**D.** Geographical external validation dataset from the National Center of Neurology and Psychiatry (NCNP). STE detector: Short-Time Energy detector. SLL: Short Line Length. HIL: Hilbert. MNI: Montreal Neurological Institute. * p < 0.05; ** p < 0.0125 derived from the DeLong test.

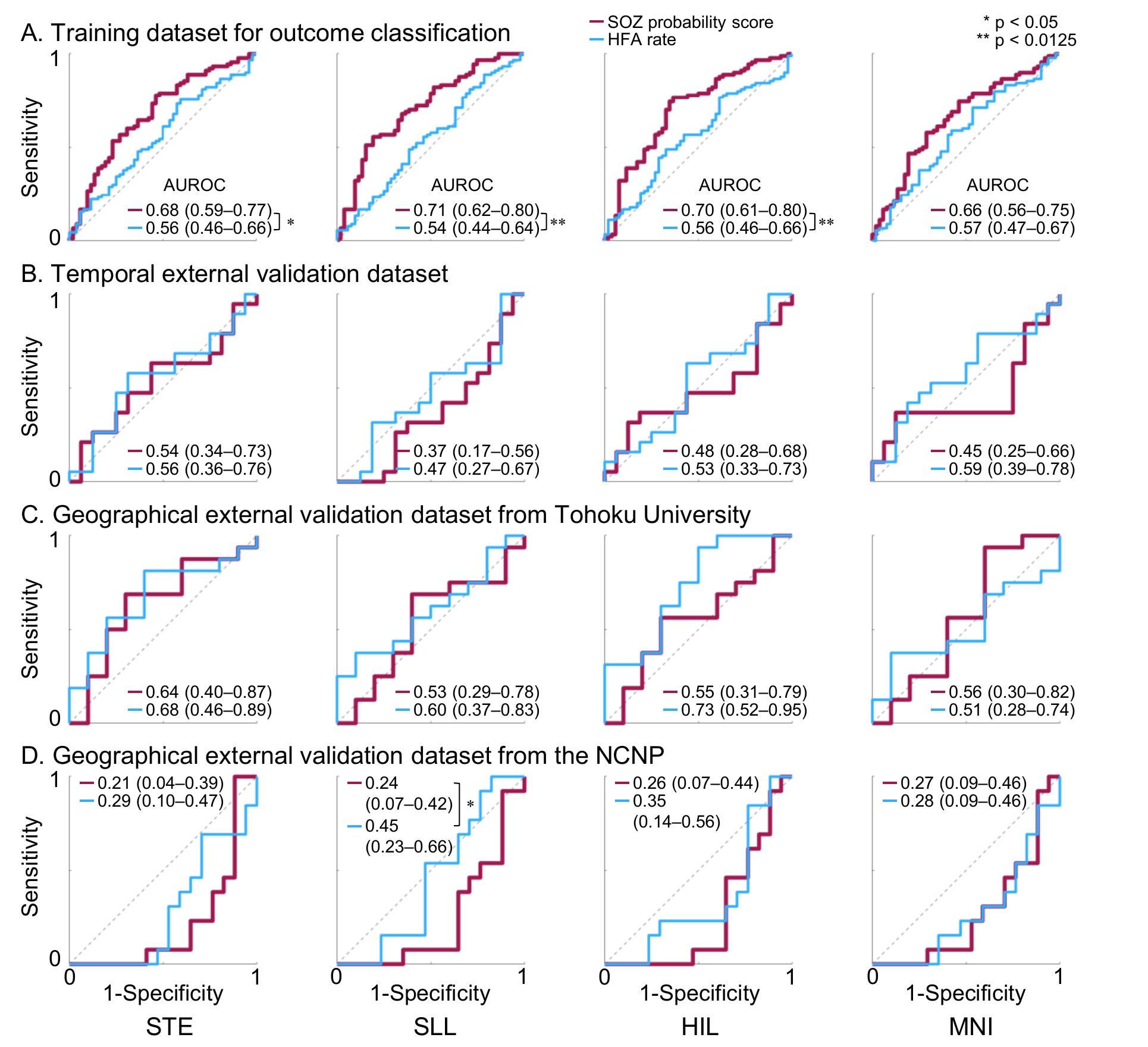

**Figure S11. Outcome prediction using ‘distinguishability statistic’ (Taylor et al., 2022).**

Receiver operating characteristic (ROC) curves present classification performance for classifying patients who achieved an ILAE class 1 outcome using the ‘seizure onset zone (SOZ) probability distinguishability statistic’ (red) and the ‘high-frequency activity (HFA) rate distinguishability statistic’ (blue). For each curve, the area under the ROC curve (AUROC) with its 95% confidence interval (95% CI) is shown.

**A.** Training dataset for outcome classification.

**B.** Temporal external validation dataset.

**C.** Geographical external validation dataset from Tohoku University.

**D.** Geographical external validation dataset from the National Center of Neurology and Psychiatry (NCNP). STE detector: Short-Time Energy detector. SLL: Short Line Length. HIL: Hilbert. MNI: Montreal Neurological Institute. * p < 0.05; ** p < 0.0125 derived from the DeLong test.
